# Should antigen-antibody rapid diagnostic tests be used to detect acute HIV infection? A systematic review and meta-analysis of diagnostic performance

**DOI:** 10.1101/2025.10.14.25338004

**Authors:** Emmanuel Fajardo, Céline Lastrucci, Pascal Jolivet, Magdalena DiChiara, Carlota Baptista da Silva, Busi Msimanga, Anita Sands, Cheryl Johnson

**Author notes:** **Corresponding author:** Cheryl Johnson, Global HIV, Tuberculosis, Hepatitis, and STI Department, World Health Organization (WHO), Avenue Appia 20, 1211 Geneva, Switzerland. **E-mail addresses of authors:** Emmanuel Fajardo, Céline Lastrucci, Pascal Jolivet, Magdalena DiChiara, Carlota Baptista da Silva, Busi Msimanga, Anita Sands, Cheryl Johnson.

## Abstract

**Introduction:** Prompt HIV diagnosis is essential to enabling effective prevention and treatment, thereby interrupting transmission and sustain epidemic control. Strategies that could diagnose individuals during acute HIV infection (AHI), while leveraging standard systems that deliver same visit results, have potential to further enhance impact. Antibody-only (Ab) rapid diagnostic tests (RDTs), which are unable to detect AHI, remain the global standard, particularly in resource-limited settings. Antigen-antibody (Ag/Ab) RDTs have been available for more than a decade, however, their use has been largely limited to high-income settings and their ability to detect AHI in resource-limited settings remains unclear.

**Methods:** We systematically searched six databases through November 2024 for studies evaluating Ag/Ab RDTs vs laboratory reference standards in individuals aged ≥18 months. Outcomes included diagnostic accuracy, patient-level outcomes, and implementation. Risk of bias was assessed with QUADAS-2, JBI Critical Appraisal and ROBINS-I tools. Diagnostic accuracy was meta-analyzed with a bivariate random-effect model; other outcomes were synthesized narratively.

**Results:** From 1,366 screened records, 53 studies from 24 countries were included. The pooled sensitivity of Ag/Ab RDTs for AHI was only 48% (95% CI: 34–62) with specificity of 97% (95% CI: 84–100). Analytical sensitivity (detection of p24 antigen) was 31% and antibody detection was 15%. Ag/Ab RDT sensitivity was higher in studies using serum/plasma specimens (66%) or in laboratory setting (54%) and compared to studies using fingerstick/capillary or venous blood (33–35%). Among oral PrEP users, Ag/Ab RDTs sensitivity was 25% (95% CI, 13–44) and specificity of 94% (95% CI, 64–99). HIV positivity varied widely across population. Uptake, feasibility and linkage were generally favourable, although challenges in resource-limited settings were noted.

**Discussion:** Ag/Ab RDTs appear to have limited ability to detect AHI, missing more than half of AHI cases, especially in resource-limited-settings and when using fingerstick/whole blood samples and implemented within PrEP programmes. Further research should evaluate cost-effectiveness, feasibility, and implementation under real-world conditions in resource-limited contexts.

**Conclusions:** Detection of AHI using Ag/Ab RDTs remains a challenge. Programmes should continue to prioritize maintaining flexible, low-cost testing strategies aligned with WHO guidance to maximize access and impact, based on their setting and context.

## 1 INTRODUCTION

Prompt and accurate HIV diagnosis is fundamental to achieving global goals to end HIV as a public health threat by 2030 [1]. HIV testing remains the essential gateway to prevention and treatment, including emerging long-acting products; ensuring that testing is accessible, reliable and responsive to evolving needs is therefore critical [2–4]. Early diagnosis, particularly during acute HIV infection (AHI), could offer an opportunity to enhance both individual and public health benefits. Immediate treatment initiation during AHI could reduce transmission and improve clinical outcomes [5].

AHI, defined by the highly infectious phase of early infection, typically within two-weeks after HIV-exposure, and may present with nonspecific or flu-like symptoms [6]. At the individual-level, HIV diagnosis during this brief window remains challenging, with recent data showing that due to declining testing volumes, and late-presentation, that the gap between HIV acquisition and diagnosis has doubled since 2017 [7]. Technologically, antibody (Ab)-only rapid diagnostic test (RDTs)—the global standard for routine HIV testing cannot reliably detect AHI and is known to miss early cases [8]. To date, reliable detection of HIV during AHI has required nucleic acid testing (NAT) techniques, or other laboratory-based Ag/Ab assays, which are costly do not provide same visit results, and are not routinely available in many settings.

While some programmatic approaches, such as outreach testing and AHI screening tools have shown to be beneficial in research settings, their real-world effectiveness and feasibility may vary [9]. Renewed attention has been drawn to the implications of missed AHI recently with efforts to scale-up pre-exposure prophylaxis (PrEP), including long-acting PrEP formulations, and addressing concerns about potential risks of HIV drug resistance and onward transmission.

HIV-1/2 p24 Ag/Ab RDTs (hereafter referred to Ag/Ab RDTs) that detect AHI could represent a potential innovation in programmatic significance as they are more compatible with the World Health Organization (WHO) guidance supporting decentralised, affordable and accessible testing with same visit diagnosis. This review examines the clinical utility of Ag/Ab RDTs, with a focus on diagnostic performance, client-level outcomes (including uptake, positivity, linkage, feasibility and acceptability, preferences, social harm) as well as systems-level evidence on resource implications and both facilitators and barriers to implementation. The analysis builds on a previous scoping review [5], and other systematic reviews [10,11] to assess whether Ag/Ab RDTs can strengthen service delivery and enhance public health impact.

## 2 METHODS

We registered the protocol in PROSPERO (CRD42024603346) and conducted the review according to PRISMA guidelines (Appendix S1) and the Cochrane Handbook for Diagnostic Test Accuracy Reviews [12,13]. Ethical review and informed consent were not required for this study, as it did not involve the collection or analysis of individual human subject data.

### 2.1 Eligibility criteria

We included studies assessing Ag/Ab RDTs for AHI in populations aged ≥18 months, regardless of antiretroviral exposure or diagnostic algorithm use. Eligible outcomes included diagnostic accuracy, uptake, positivity, linkage to care, feasibility, acceptability, values and preferences, resource use, social harms and barriers and facilitators to implementation. Reviews, case reports, manufacturer documents, opinion papers, and proof-of-concept studies were excluded.

### 2.2 Search strategy and selection

We searched ClinicalTrials.gov, Cochrane Library, PubMed, Medline, Scopus, and Web of Science (October–November 2024) using database-specific combinations of key words and MeSH terms (e.g., “acute HIV,” “antigen-antibody,” “rapid diagnostic tests”). Searches were restricted to January 2010–November 2024 to reflect commercial test availability. No restrictions on language, country or setting were applied; non-English studies were translated with DeepL. Additional records were identified through reference screening. Full search strategies are provided in (Appendix S2). Two independent reviewers (EF, CL) independently screened titles/abstracts and extracted data using standardized forms. Discrepancies were resolved by consensus or third-party adjudication.

### 2.3 Risk of bias assessment

Diagnostic accuracy studies were appraised with QUADAS-2, evaluating the risk of bias and applicability across four domains [14]. Patient-level impact and implementation studies were assessed with the appropriate Johanna Brigg’s Institute (JBI) Critical Appraisal Tool and ROBINS-I tool [15]. Results were summarized in tables and figures.

### 2.4 Data analysis

#### 2.4.1 Diagnostic accuracy

We included studies comparing Ag/Ab RDTs to serological and/or molecular laboratory-based assays. AHI was defined as a positive p24 antigen or HIV RNA test with seronegative or discordant antibody results. We excluded studies using nonclinical specimens or lacking raw data to calculate sensitivity and specificity.

Accuracy measures were pooled by reference standard, with sensitivity and specificity reported alongside 95% CIs. Inconclusive results were excluded from pooled analysis but reported separately. Clinical utility of AHI detection was assessed as the relative increase in case detection when comparing Ag/Ab RDTs with antibody-only RDTs. Reference standards were categorized as NAT-based or p24 Ag-based for subgroup analysis.

Analyses were conducted in StataNow 19.5 (StataCorp, College Station, TX). Stata “metadta” was used to produce pooled analysis and forest plots [16]. Bayesian re-analysis was performed in R to verify robustness.

#### 2.4.2 Patient-level and implementation outcomes

In addition to evaluating the clinical utility of AHI detection, we analyzed patient-level outcomes descriptively, including test uptake, overall positivity and antigen-only positivity, linkage to confirmatory testing, linkage to ART initiation, and turnaround time to treatment. These outcomes are reported as percentages where applicable. Broader implementation outcomes – feasibility, acceptability, values/preferences, social harms, resource use and barriers and facilitators to implementation –were summarized narratively.

## 3 RESULTS

The search identified 2,288 records; after removing duplicates, 1,366 unique records were screened, and 53 studies were included (Figure 1). Of these, 31 were diagnostic test accuracy studies [17–47], 12 evaluated patient-level impact [43, 48, 50–52, 54, 60, 62–64, 67, 68], 14 addressed feasibility [43, 48, 50–52, 54, 59–60, 62–64, 66–68], nine explored values and preferences [49, 51, 53–56, 61, 68, 69], three examined resource use [57, 58, 65], and 19 reported barriers and facilitators to implementation. Study categories were not mutually exclusive. A detailed summary of outcomes reported by each study is available in (Table S1).

**Figure 1.**
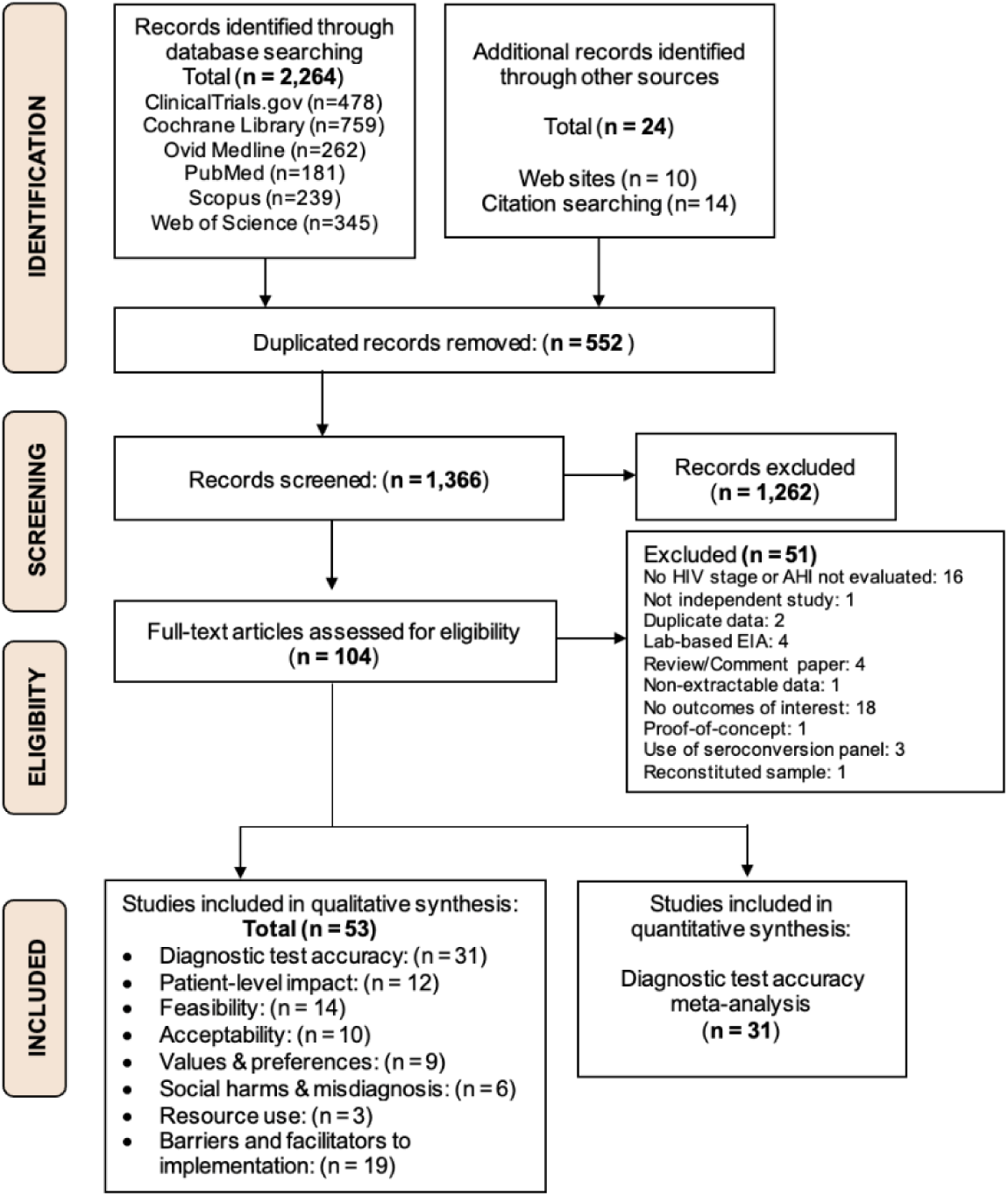
PRISMA flowchart of included and excluded records.

### 3.1 Study characteristics

Thirty-one studies reported on diagnostic accuracy and characteristics are summarized in Table 1. The 31 studies, spanning all five WHO regions were primarily in health facilities (53% laboratories, 33% clinics). Populations included individuals at high ongoing HIV risk (40%), those previously diagnosed with HIV (37%), PrEP users (10%), hospital patients (7%), the general population (3%) and women (3%). All studies evaluated adults; none assessed infants or children. Only six studies (16%) used fingerstick or venous blood with most relying on composite reference standards to define infection status. Across studies, a total of 7,184 specimens were analyzed, with sample sizes ranging from 5 to 1,494; nearly two-thirds (63%) involved fewer than 100 specimens, typically in laboratory-based case-control studies. Details on RDT brands and testing operators are provided in (Table S2).

**Table 1.**
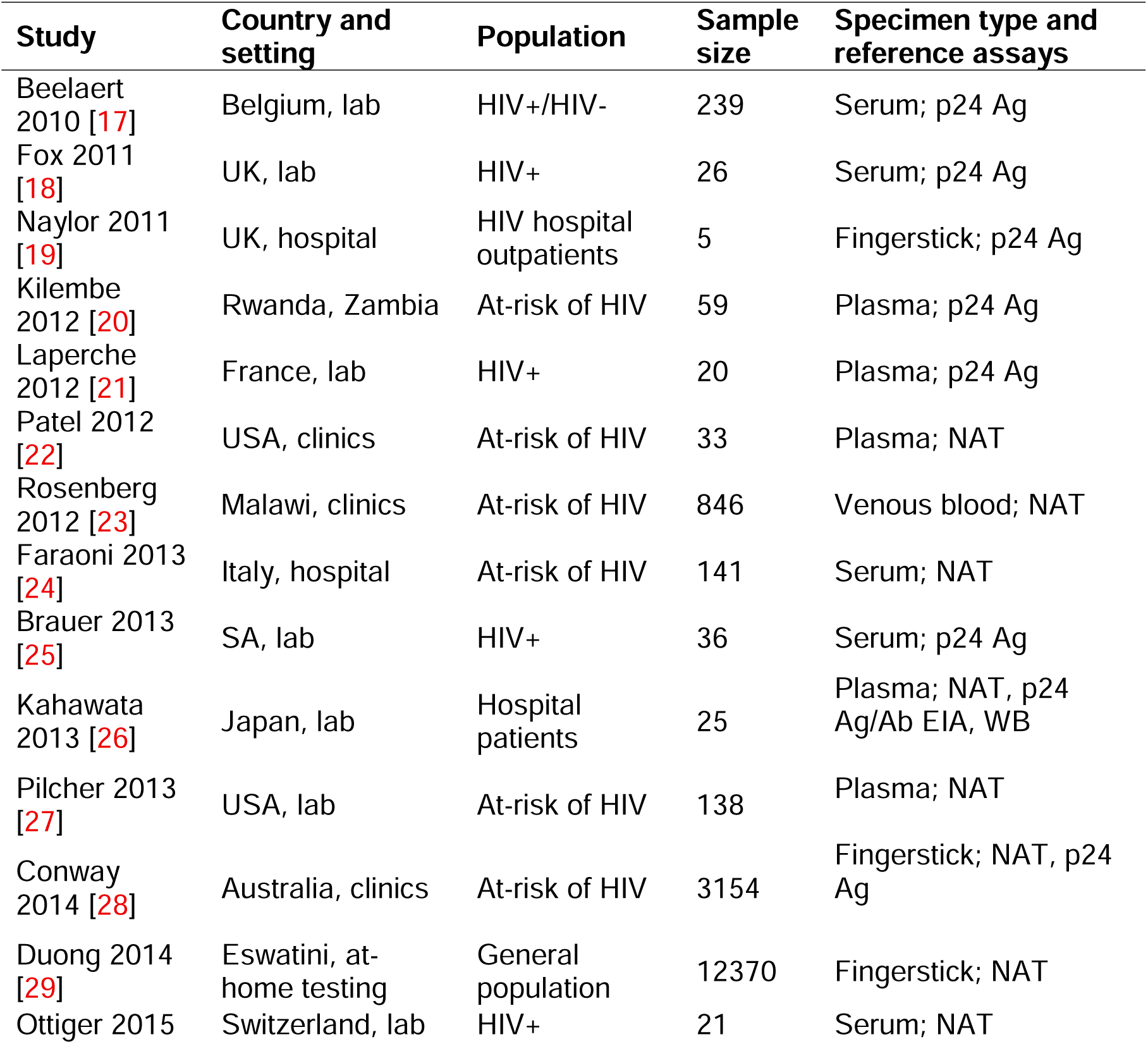
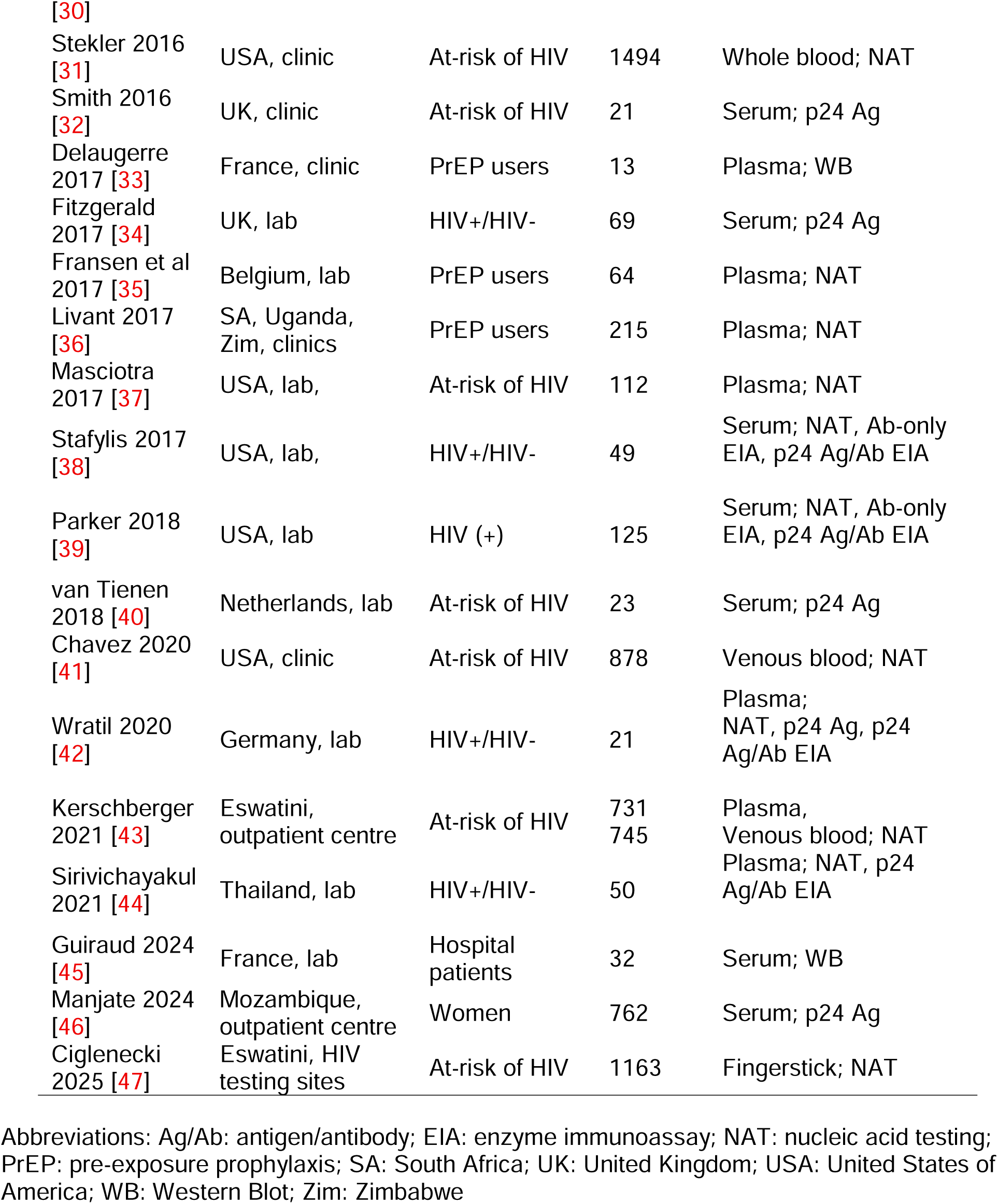
Key characteristics of diagnostic accuracy studies.

An additional 23 studies reported on client-level and implementation outcomes [48–69], Further details and study characteristics are provided in (Table S3).

### 3.2 Risk of bias assessment

Study quality was assesses using QUADAS-2 (Table S4). Most diagnostic accuracy studies were judged to have a low risk of bias for the index test, reference standard, and flow/timing domains. However, risk of bias was frequently high for patient selection, reflecting the predominance of case-control designs and non-consecutive sampling. Concerns regarding applicability were generally low across domains. See (Table S4) for full assessment.

Study quality for thirteen non-diagnostic accuracy studies was evaluated using the JBI Critical Appraisal Tool, and three studies were assessed with the ROBINS-I tool. All cross-sectional studies demonstrated a low risk of bias (Table S5). Among the three cohort studies, two had a low risk of bias, while one was judged to have a high risk due to missing data, outcome measurement issues, and selective reporting (Table S6).

### 3.3 Diagnostic accuracy

A meta-analysis of 31 studies, representing 38 reports, had a pooled diagnostic sensitivity and specificity of Ag/Ab RDTs for AHI of 48% (95% CI: 34–62) and 97% (95% CI: 84–100), respectively, with considerable heterogeneity across studies (Figure 2). Pooled sensitivity and specificity for p24 antigen detection varied widely (31%, 95% CI: 19–45 to 99%, 95% CI: 97–100), with considerable heterogeneity across studies (Figure 3).

**Figure 2.**
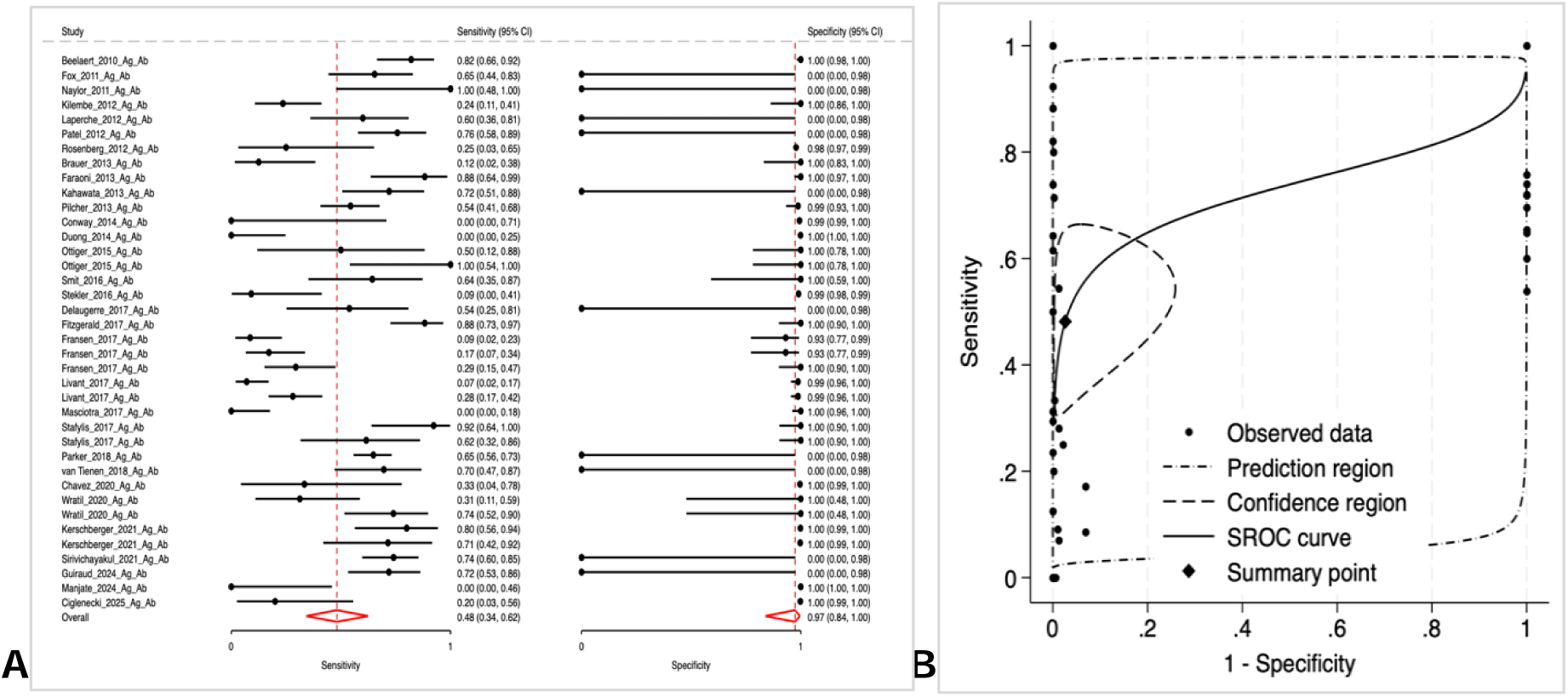
Diagnostic accuracy of Ag/Ab RDTs. (A) Forest plot of sensitivity and specificity from individual studies with 95% confidence intervals; pooled estimates are shown as red diamonds. (B). Summary receiver operating characteristics (SROC) curve with 95% confidence and prediction regions.

**Figure 3.**
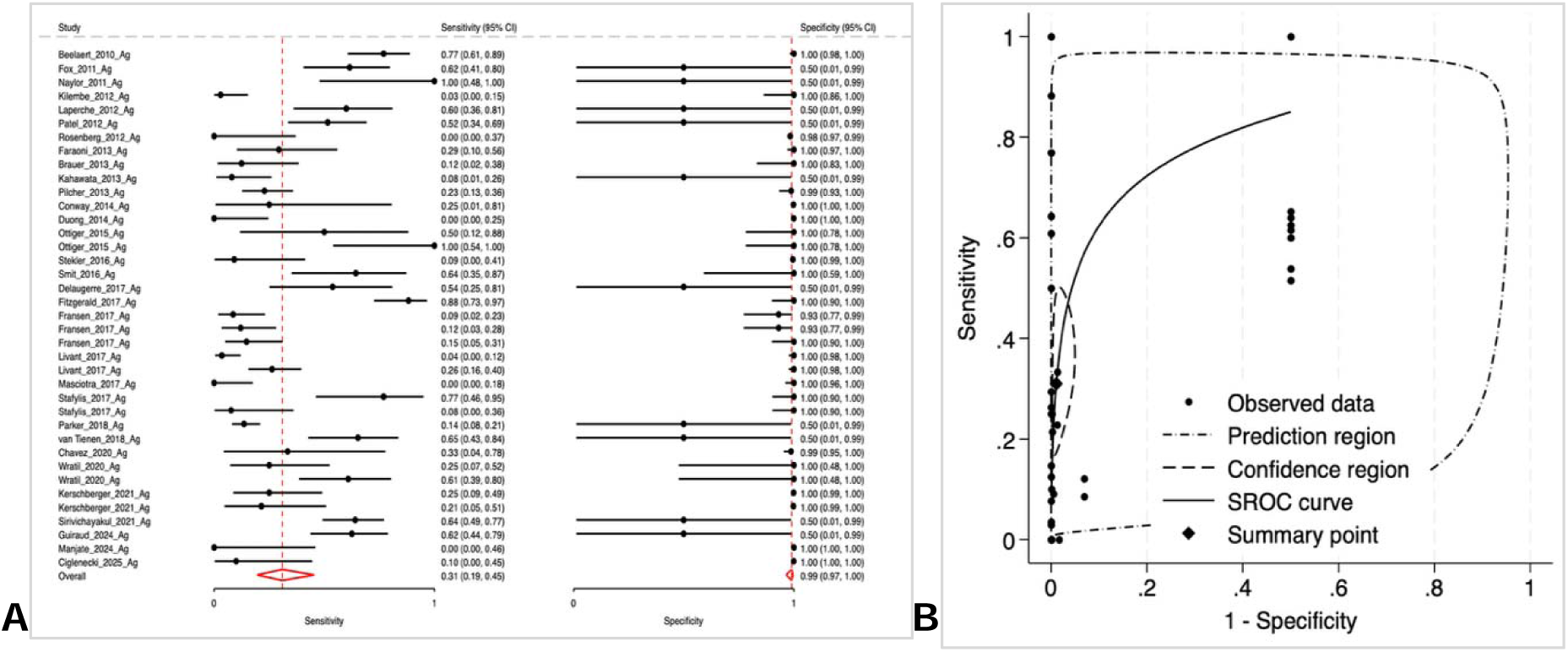
Diagnostic accuracy of Ag/Ab RDTs (p24 Ag detection). (A) Forest plot of sensitivity and specificity from individual studies with 95% confidence intervals; pooled estimates are shown as red diamonds. (B). Summary receiver operating characteristics (SROC) curve with 95% confidence and prediction regions.

The pooled estimates for HIV-antibody detection also ranged widely (15% sensitivity, 95% CI: 10–24 to 99% specificity, 95% CI: 96–100), with substantial heterogeneity (Figure 4).

**Figure 4.**
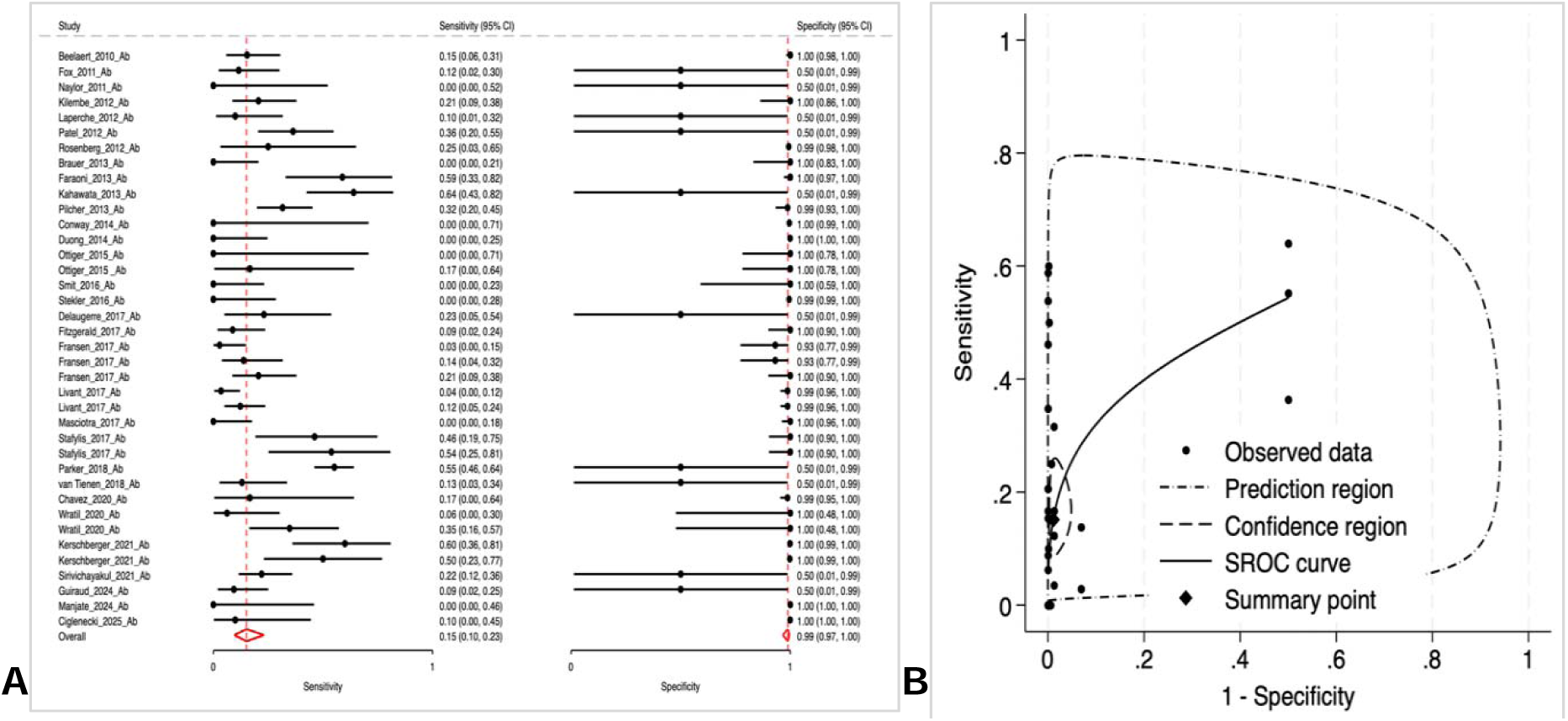
Diagnostic accuracy of Ag/Ab RDTs (antibody detection). (A) Forest plot of sensitivity and specificity from individual studies with 95% confidence intervals; pooled estimates are shown as red diamonds. (B). Summary receiver operating characteristics (SROC) curve with 95% confidence and prediction regions.

In subgroup analysis (Table 2), sensitivity was higher in case-control studies (61%), European countries (70%), serum-based testing (66%), laboratory settings (54%), when using p24 Ag reference assays (58%), and among people living with HIV who had previously been diagnosed (70%). The accuracy of Ag/Ab RDTs in capillary/fingerstick and venous whole blood specimens was lower compared to serum and plasma, with sensitivities of 35% and 33%, respectively, while specificities remained above 99%. Among oral PrEP users, sensitivity and specificity dropped to 25% (95% CI, 13–44) and 94% (95% CI, 64–99).

**Table 2.** Subgroup analysis of pooled accuracy estimates for Ag/Ab RDTs.

| WHO region | p24 Ag/Ab combined |  | p24 Ag detection |  | Ab detection |  |
| --- | --- | --- | --- | --- | --- | --- |
|  | Sensitivity (95% CI) | Specificity (95% CI) | Sensitivity (95% CI) | Specificity (95% CI) | Sensitivity (95% CI) | Specificity (95% CI) |
| African | 17%<br>(5–44) | 100%<br>(99–100) | 12%<br>(8–19) | 100%<br>(99–100) | 13%<br>(6–31) | 100%<br>(99–100) |
| The Americas | 39%<br>(14–71) | 99%<br>(92–100) | 21%<br>(11–37) | 99%<br>(97–100) | 26%<br>(12–47) | 99%<br>(99–100) |
| European | 70%<br>(56–81) | 98%<br>(83–100) | 61%<br>(44–75) | 95%<br>(83–99) | 14%<br>(9–26) | 95%<br>(77–99) |
| South-East Asia | – | – | – | – | – | – |
| Western Pacific | – | – | – | – | – | – |
| **Study setting** |  |  |  |  |  |  |
| Laboratory | 54%<br>(38–69) | 97%<br>(92–99) | 34%<br>(20–50) | 98%<br>(92–99) | 19%<br>(12–28) | 98%<br>(92–100) |
| Non-laboratory | 40%<br>(14–73) | 98%<br>(93–100) | 34%<br>(19–53) | 98%<br>(90–99) | 13%<br>(5–28) | 98%<br>(92–100) |

| **Study design** |  |  |  |  |  |  |
| --- | --- | --- | --- | --- | --- | --- |
| Case-control | 61%<br>(50–72) | 97%<br>(89–99) | 42%<br>(29–57) | 95%<br>(86–98) | 21%<br>(14–30) | 96%<br>(85–99) |
| Cross-sectional | 9%<br>(0–69) | 100%<br>(98–100) | 17%<br>(7–37) | 100%<br>(98–100) | 10%<br>(3–29) | 100%<br>(99–100) |
| **Population** |  |  |  |  |  |  |
| HIV-infected | 70%<br>(59–79) | 97%<br>(78–100) | 60%<br>(43–74) | 96%<br>(82–99) | 17%<br>(9–29) | 96%<br>(77–99) |
| At-risk of HIV | 29%<br>(11–57) | 99%<br>(98–100) | 18%<br>(10–32) | 99%<br>(97–100) | 18%<br>(9–31) | 100%<br>(99–100) |
| PrEP users | 25%<br>(13–44) | 94%<br>(64–99) | 21%<br>(9–43) | 97%<br>(44–100) | 12%<br>(7–22) | 96%<br>(78–99) |
| **Specimen type** |  |  |  |  |  |  |
| Serum | 66%<br>(51–78) | 99%<br>(87–100) | 46%<br>(26–68) | 97%<br>(85–99) | 17%<br>(8–30) | 95%<br>(82–99) |
| Plasma | 43%<br>(23–67) | 98%<br>(87–100) | 24%<br>(13–39) | 98%<br>(84–100) | 23%<br>(13–38) | 96%<br>(82–99) |
| Venous blood | 33%<br>(13–63) | 99%<br>(98–99) | 17%<br>(8–33) | 99%<br>(98–100) | 19%<br>(5–50) | 99%<br>(99–100) |
| Fingerstick blood | 35%<br>(1–96) | 99%<br>(93–99) | 24%<br>(4–71) | 99%<br>(95–100) | 1%<br>(0–66) | 99%<br>(99–100) |
| **Reference standard** |  |  |  |  |  |  |
| p24 Ag | 58%<br>(33–79) | 100%<br>(77–100) | 45%<br>(17–77) | 99%<br>(86–100) | 11%<br>(7–16) | 99%<br>(81–100) |
| NAT | 35%<br>(14–64) | 99%<br>(98–100) | 21%<br>(9–42) | 99%<br>(98–100) | 15%<br>(6–31) | 99%<br>(99–100) |

Overall, there was a high-level of variation in the total number of additional AHI cases identified by the antibody component of Ag/Ab RDTs compared with Ab-only RDTs [22,27,33,35,36,45] (Figure 5).

**Figure 5.**
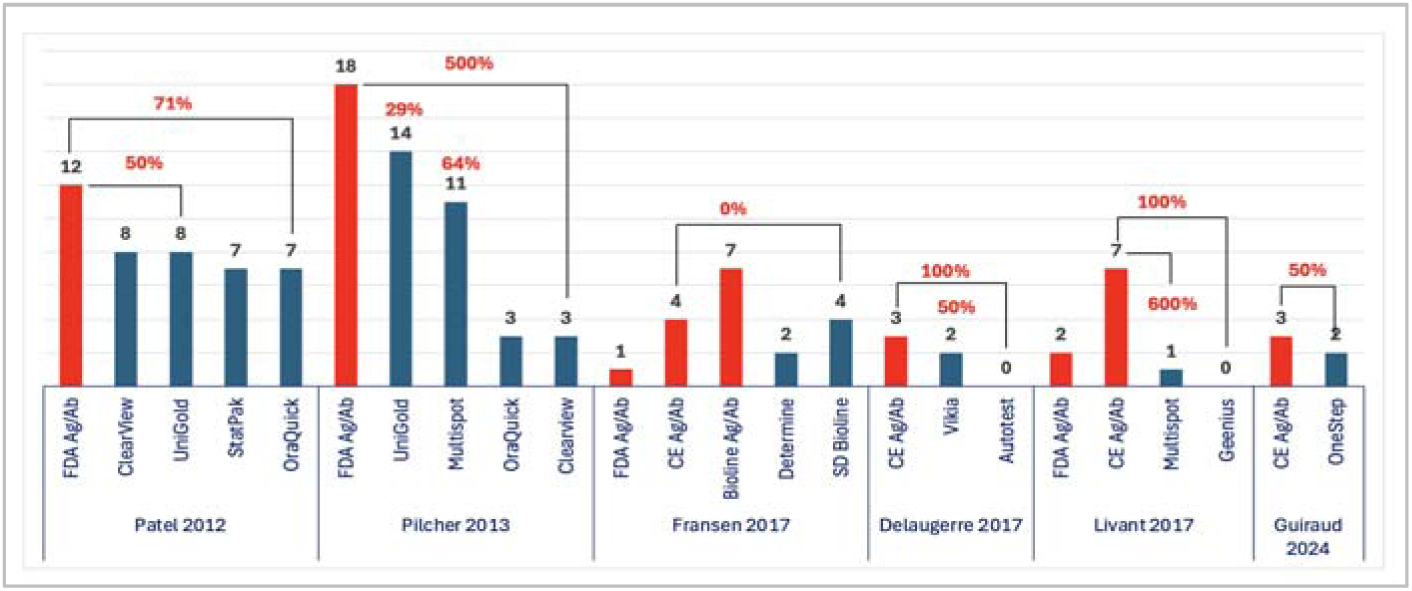
Relative increase in AHI cases detected by the antibody component of Ag/Ab RDTs compared with Ab-only RDTs.

### 3.4 Client-level impact outcomes

#### 3.4.1 Uptake and positivity

Four observational studies assessed uptake of Ag/Ab RDTs, with high with rates reported among vulnerable and key populations in Belgium (78%), Argentina (100%) and Spain (100%) [52,62,67], but low initial uptake (14.3%) in primary healthcare clinics in MSM in Australia, which improved over time [51]. Twelve studies reported wide variation in overall test positivity rates (mostly antibody-based) ranging from 0.77% in general populations in the USA [63], 1.3% – 7.7% among at-risk individuals and key populations [43,48,50–52,54,64,67], and 8.1% in antenatal clinics in Kenya [68], up to 19.6% among transgender women in Argentina [62] and high rates in key populations using partner notification services in Vietnam [60]. Nine studies documented p24 antigen-only reactivity: four reported 0% (UK, Australia, Argentina and Spain [48,51,62,67], while positivity reached 7% in the USA [63], 10% in pregnant women in Kenya [68], 11% among key populations in Vietnam [60], 20% in at-risk individuals in Eswatini [43], and 31% among MSM in a community-based clinic in Spain [64].

#### 3.4.2 Linkage to confirmatory testing and treatment

Across studies linkage to confirmatory testing were generally high, as with other HIV testing studies. Ab-reactive results (established HIV) achieved 99%–100% in seven studies [48,50–52,54,62,67]. In four studies, all Ag-reactive cases (100%) were reportedly confirmed, either later on site or through referrals off-site [43,60,63,64].

Turnaround times for confirmatory testing varied from 1–7 day in high-income contexts only [55,56,63]. Treatment initiation after confirmed diagnosis was high, with rates of 86-100% [48,50,52,54,63,64,67], primarily in high-income countries. However, in resource-limited settings, such as Eswatini, delays were common with only 72% initiating treatment within one month, 37% initiating within one week and 13% initiating same day [43]. In Vietnam, barriers such as patient readiness and delays in confirmatory testing also hindered timely treatment [60].

#### 3.4.3 Additional implementation outcomes

Nineteen studies documented facilitators and barriers to implementation [43, 48–57, 59–64, 66–68]. Facilitators included user-friendliness, convenience, and rapid results. high acceptability among clients and healthcare providers due to privacy, confidentiality, and anonymity. Low-cost testing with demonstrated cost-effectiveness [57–59, 67, 68], including willingness to pay if unfunded [55]. Barriers included interpretation challenges for inexperienced operators, particularly with three-test-line results, highlighting the need for robust training [63]. A comprehensive list of facilitators and barriers is provided in Table 3.

**Table 3.**
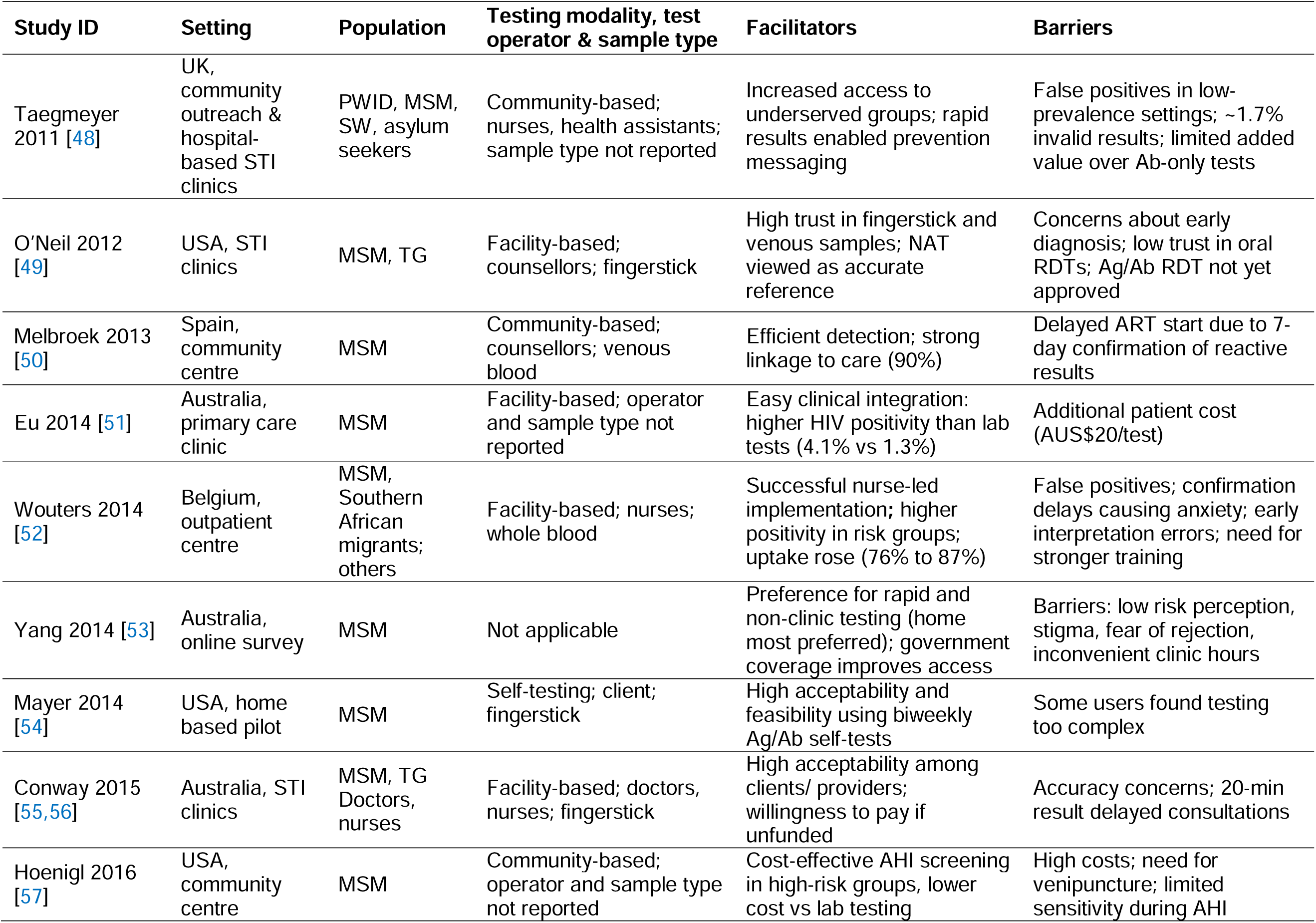
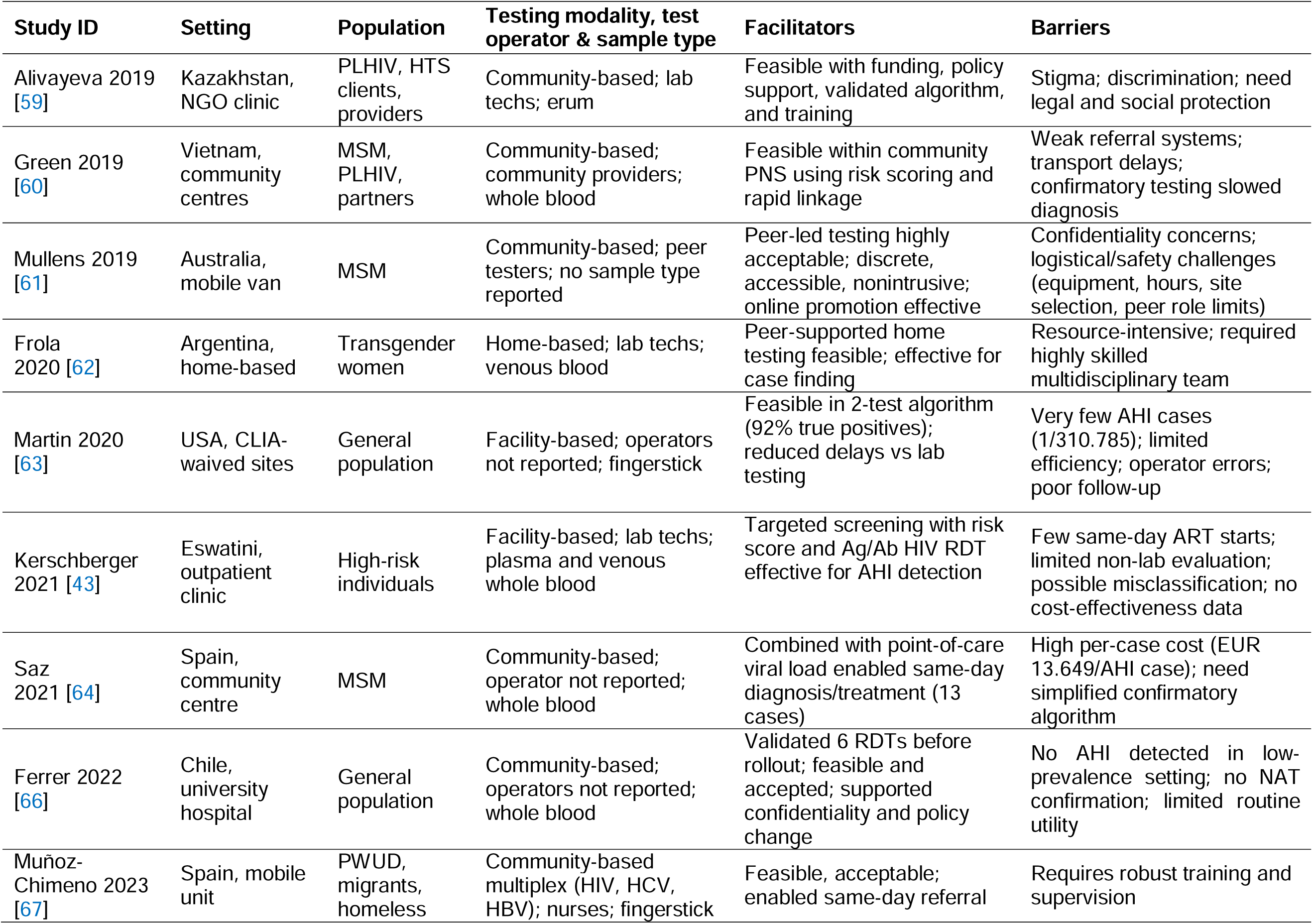
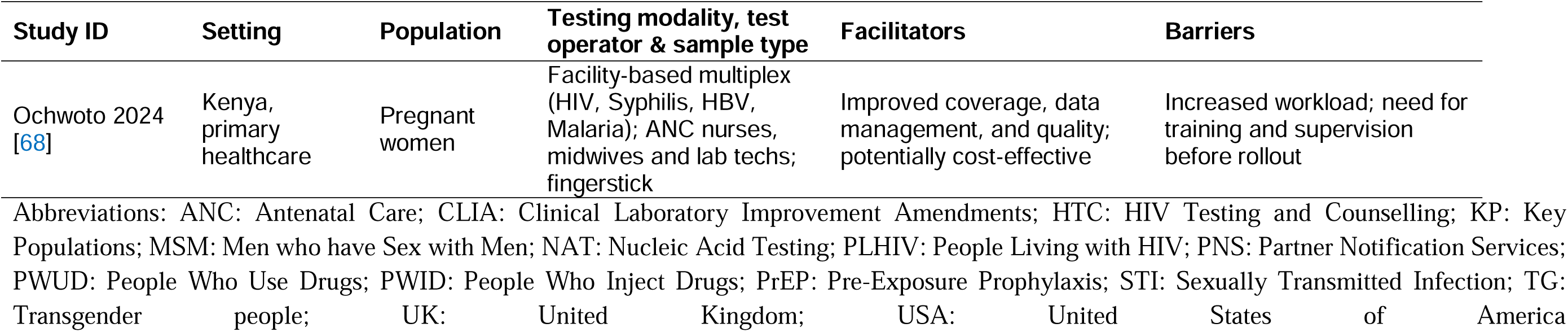
Studies reporting barriers and facilitators to implementation.

Fourteen studies suggested Ag/Ab RDTs could be acceptable and feasible to implement, but there was considerable variability by setting, test provider, and population. Reported social harms were rare, though false positives occurred, especially during early implementation, highlighting the importance of training and quality assurance. Resource use information was limited to high-income settings, but suggested Ag/Ab RDTs could be cost-effective compared to alternatives, particularly in higher prevalence or high-risk populations. Key facilitators to success and buy-in included implementation that was user-friendliness, discreet, able to track reactive results in real time; however, barriers to implementation included challenges due to limited training, operator inexperience, lack of interchangeability with other products, stigma, legal and financial constraints, and confirmatory testing delays. Further details on implementation related outcomes are provided in (Appendix S3).

## 4 DISCUSSION

The findings from this review, which comprehensively evaluated the diagnostic accuracy of Ag/Ab RDTs across multiple domains, highlights that Ag/Ab RDTs have a low to moderate ability to detect AHI (48%, 95% CI: 34–62) and that results per country, context and population may vary widely. Performance using capillary/fingerstick or venous blood samples, and outside of laboratory settings, had low sensitivity during AHI; 35% and 33%, respectively. Based on current data summarized by this review, Ag/Ab RDTs appear to have several limitations and variable performance in terms of their ability to detect AHI within routine HIV testing in low-and middle-income countries, particularly those relying on decentralised routine testing outside laboratories [23,28,29,31,41,47]. These findings are consistent with earlier studies and reviews that compared HIV Ag/Ab RDTs with Ab-only RDTs [22,27,33,35,36,45].

This review also highlighted that while there is interest in the role Ag/Ab RDTs could play in PrEP implementation, test performance during AHI among PrEP users was poor, with pooled sensitivity and specificity of 23% (95% CI: 16–33) and 87% (95% CI: 66–96), respectively. This remains lower than established performance of laboratory-based Ag/Ab assays and NAT. Encouragingly, however, in the most recent update to the WHO guidelines evidence showed that standard Ab-only RDTs, as well as self-tests, are sufficient for PrEP and LAI-PrEP which is important to continue access to these important prevention tools in resource-limited sites [70,71]. A recent modelling study highlighted that existing Ab-only RDTs, as well as self-tests, with even yearly testing for LAI-PrEP would not have a significant impact in incidence or drug resistance [72,73]. Similarly, results from Brazil’s implementation of CAB-LA show self-tests performed well and were sufficiently accurate [74].

Despite the limitations related to AHI detection in a routine decentralized testing service, quality-assured Ag/Ab RDTs may have benefits, as indicated in select studies, when implemented in settings with limited access to sensitive assays for blood screening and safety [75] or as part of highly targeted case-finding and outreach efforts. Implementation studies highlighted that given available systems and resources, Ag/Ab RDTs, could offer value and be cost-effective if highly targeted [65]. Even with these potential possibilities, however, efforts to place these tools and achieve impact could be challenging. With resources, and staffing, declining, there are challenges with introducing tests that may also increase the cost of training and limit the ability to have flexible or interchangeable algorithms. Any country or programme considering Ag/Ab RDTs will need to ensure they have the resources to support the potentially higher costs, as well as transition costs, given shrinking testing capacity globally. WHO’s tool kit, low-cost diagnostic strategy and prioritization guide are important resources that may be helpful to consider in this uncertain environment as the promote low cost and flexible product selection based on local needs [76].

Cost-effectiveness analysis, as well as budget impact analysis, with real implementation data was not available in any resource-limited setting and is a data gap that should be addressed. Understanding the benefits, and costs, of adopting other assays such as multiplex tests need to be considered to ensure investment is going toward interventions that have greatest value for money, and which will have substantial public health impact.

This review has several strengths: it integrates pooled diagnostic accuracy estimates with patient-level and implementation outcomes, drawing from 53 studies across 25 countries. Its breadth enhances generalizability and relevance, while identifying implementation barriers and research gaps. By breaking down evidence by geography, population and sample type additional insights for real world settings can also be derived.

Despite these strengths, a critical limitation was the publication bias with most evidence being classified as low certainty due to the large number of case-control studies included [77]. First, most available evidence originates from high-resource settings, leaving important gaps in low-and middle-income countries. High-quality, real-world evaluations on the public health impact, and clinical utility, of Ag/Ab HIV RDTs in routine HIV testing services in resource-limited settings remain important research gaps to fill. Additional gaps identified also included the inability to evaluate test performance in children and the high-level of heterogeneity in how studies defined and measured AHI. Further, while PrEP users were included, because of the timing of the review and the diversity of products entering the market, our results may also not be fully representative of all forms of PrEP being implemented. Although we extracted, standardized and recalculated data as much as possible there is a risk that we included individuals without AHI in our estimates, or missed studies due to reporting not being sufficiently disaggregated. The review also focuses on detection of AHI and our findings focus on the additional benefits, and do not provide a full review of all aspects of diagnostic performance. Changes in the commercially available products may also affect results as this review is focused on the performance of a broad class of Ag/Ab rapid tests, and how they may perform and how their general use may affect programmes, and is not focused on specific brands.

Lastly, while we discuss the importance of data on cost-effectiveness and resource use, there was insufficient data available, specifically from resource-limited settings. This is critical moment with substantial uncertainties on future investment and funding for global health. Costs are shifting and if Ag/Ab RDTs costs decrease and become similar or lower cost when compared to alternative Ab-only RDTs this would be an important consideration. As with all technologies, before introducing new products, it remains critical that countries and funders understand the short-, medium- and long-term cost implications of adopting Ag/Ab RDTs.

More data on the real-world feasibility of using Ag/Ab RDTs in routine testing settings with minimal resource requirements, including client level-impact, particularly studies evaluating the impact of streamlined pathways for rapid confirmation of AHI (including standardised case definition) and ART initiation, would be beneficial. Additional economic analysis along with values and preferences and social harms would also strengthen the evidence base and address knowledge gaps.

## 5 CONCLUSIONS

Prompt and accurate HIV diagnosis remains essential to ending the epidemic. HIV RDTs remain critical to the response. The added value of Ag/Ab RDTs remains unclear due to limited and variable sensitivity during AHI—particularly when using fingerstick/capillary blood specimens.

WHO guidance remains the same and there is no evidence suggesting Ag/Ab RDTs should be prioritized within routine testing services, including for PrEP delivery. Rather, given the shortages in staff and funding it is important countries embrace WHO’s guidance on ensuring country flexibilities and work toward adopting low-cost commodities to maintain testing coverage where it is needed most.

Programmes in resource-limited settings must continue to carefully balance the risks and benefits of adopting different technologies which may increase complexity or costs, and factor in potential competing priorities (e.g. multiplex testing and self-testing.

## AUTHORS’ CONTRIBUTIONS

Conceptualization: CJ, AS, EF, CL; Methodology: EF, CJ; Formal analysis: EF, CL; Data curation: EF; Investigation: EF, CL, CJ, AS; Writing – original draft: EF, CJ; Writing – review & editing: EF, CL, PJ, MD, CBS, BS, AS, CJ; Visualization: EF; Supervision: CJ, MD; Project administration: CJ, MD; Funding acquisition: CJ

## ACKNOWLEDGEMENTS

We sincerely thank Olanrewaju Ayodele Edun for independently validating our results by rerunning the bivariate random-effect models in R using the Bayesian approach for diagnostic accuracy meta-analysis, and for his invaluable statistical guidance throughout this study.

## DISCLAIMER

The opinion and views expressed in this article are those of the authors and do not necessarily reflect the official position, policies, or decisions of the World Health Organization.

## DATA AVAILABILITY STATEMENT

All data underlying the findings of this study are included in the article and its supporting information files. Extracted datasets and analysis files are available from the corresponding author upon reasonable request.

## SUPPORTING INFORMATION

Additional information may be found under the Supporting Information tab for this article.

**Appendix S1.** PRISMA Checklist

**Appendix S2.** Full search strategies

**Appendix S3.** Feasibility, acceptability, values and preferences, social harms, and resource use.

**Table S1.** Outcomes by study and summary findings

**Table S2.** RDT brands and testing operators by study

**Table S3.** Characteristics of 23 studies assessing patient-level impact and implementation outcomes

**Table S4.** QUADAS-2 risk of bias assessment

**Table S5**. JBI risk of bias assessment

**Table S6.** ROBINS-I risk of bias assessment

## REFERENCES

1. Global health sector strategies on, respectively, HIV, viral hepatitis and sexually transmitted infections for the period 2022-2030. Geneva: World Health Organization; 2022. Available: https://iris.who.int/bitstream/handle/10665/360348/9789240053779-eng.pdf?sequence=1

2. Consolidated guidelines on differentiated HIV testing services. Geneva: World Health Organization; 2024. Available: https://www.who.int/publications/i/item/9789240096394

3. Guidelines on long-acting injectable cabotegravir for HIV prevention. Geneva: World Health Organization; 2022. Available: https://www.who.int/publications/i/item/9789240054097

4. Guidelines on lenacapavir for HIV prevention and testing strategies for long-acting injectable pre-exposure prophylaxis (PrEP). Geneva: World Health Organization; 2025. Available from: https://iris.who.int/bitstream/handle/10665/381892/9789240111608-eng.pdf?sequence=1

5. Rutstein SE, Ananworanich J, Fidler S, et al. Clinical and public health implications of acute and early HIV detection and treatment: a scoping review. J Int AIDS Soc. 2017 Jun 28;20(1):21579.

6. Bottone PD, Bartlett AH. Diagnosing Acute HIV Infection. Pediatr Ann. 2017;46(2):e47–e50.

7. Hall M, Golubchik T, Bonsall D, Abeler-Dörner L, Limbada M, Kosloff B, et al; HPTN 071 (PopART) Phylogenetics protocol team; PANGEA consortium. Demographics of sources of HIV-1 transmission in Zambia: a molecular epidemiology analysis in the HPTN 071 PopART study. Lancet Microbe. 2024 Jan;5(1):e62–e71.

8. Adetunji AA, Adewumi MO, Michael OS, Fayemiwo SA, Ogunniyi A, Taiwo BO. Rapid HIV Antigen-Antibody Assays and Detection of Acute HIV Infection in Sub-Saharan Africa. Am J Trop Med Hyg. 2019;101(2):285–286.

9. Ong JJ, Coulthard K, Quinn C, Tang MJ, Huynh T, Jamil MS, Baggaley R, Johnson C. Risk-Based Screening Tools to Optimise HIV Testing Services: a Systematic Review. Curr HIV/AIDS Rep. 2022 Apr;19(2):154–165.

10. Lewis JM, Macpherson P, Adams ER, Ochodo E, Sands A, Taegtmeyer M. Field accuracy of fourth-generation rapid diagnostic tests for acute HIV-1: a systematic review. AIDS. 2015;29(18):2465–2471.

11. Smallwood M, Vijh R, Nauche B, Lebouché B, Joseph L, Pant Pai N. Evaluation of a rapid point of care test for detecting acute and established HIV infection and examining the role of study quality on diagnostic accuracy: A Bayesian meta-Analysis. PLoS One. 2016;11(2):e0149592.

12. McInnes MDF, Moher D, Thombs BD, McGrath TA, Bossuyt PM, et al. Preferred Reporting Items for a Systematic Review and Meta-analysis of Diagnostic Test Accuracy Studies: The PRISMA-DTA Statement [published correction appears in JAMA. 2019 Nov 26;322(20):2026.

13. Deeks JJ, Bossuyt PM, Leeflang MM, Takwoingi Y (editors). Cochrane Handbook for Systematic Reviews of Diagnostic Test Accuracy. Version 2.0 (updated July 2023). Cochrane, 2023. Available: https://training.cochrane.org/handbook-diagnostic-test-accuracy/current.

14. Whiting PF, Rutjes AWS, Westwood ME, Mallett S, Deeks JJ, Reitsma JB, et al. QUADAS-2: a revised tool for the quality assessment of diagnostic accuracy studies. Ann Intern Med. 2011 Oct 18; 155(8):529–36.

15. Sterne JAC, Hernan MA, Reeves BC, Savović J, Berkman ND, Viswanathan M, et al. ROBINS-I: a tool for assessing risk of bias in non-randomised studies of interventions. BMJ. 2016 Oct 12; 355:i4919.

16. Nyaga VN, Arbyn M. Metadta: a Stata command for meta-analysis and meta-regression of diagnostic test accuracy data - a tutorial. Arch Public Health. 2022 Mar 29;80(1):95. Erratum in: Arch Public Health. 2022 Sep 27;80(1):216

17. Beelaert G, Fransen K. Evaluation of a rapid and simple fourth-generation HIV screening assay for qualitative detection of HIV p24 antigen and/or antibodies to HIV-1 and HIV-2. J Virol Methods. 2010 Sep;168(1-2):218–22.

18. Fox J, Dunn H, O’Shea S. Low rates of p24 antigen detection using a fourth-generation point of care HIV test. Sex Transm Infect. 2011 Mar;87(2):178–9.

19. Naylor E, Axten D, Makia F, Tong C, White J, Fox J. Fourth generation point of care testing for HIV: validation in an HIV-positive population. Sex Transm Infect. 2011 Jun;87(4):311.

20. Kilembe W, Keeling M, Karita E, Lakhi S, Chetty P, Price MA, Makkan H, Latka M, Likoti M, Ilukui K, Hurlston M, Allen S, Stevens G, Hunter E. Failure of a novel, rapid antigen and antibody combination test to detect antigen-positive HIV infection in African adults with early HIV infection. PLoS One. 2012;7(6):e37154.

21. Laperche S, Leballais L, Ly TD, Plantier JC. Failures in the detection of HIV p24 antigen with the Determine HIV-1/2 Ag/Ab Combo rapid test. J Infect Dis. 2012 Dec 15;206(12):1946–7; author reply 1949-50.

22. Patel P, Bennett B, Sullivan T, Parker MM, Heffelfinger JD, Sullivan PS; CDC AHI Study Group. Rapid HIV screening: missed opportunities for HIV diagnosis and prevention. J Clin Virol. 2012 May;54(1):42–7.

23. Rosenberg NE, Kamanga G, Phiri S, Nsona D, Pettifor A, Rutstein SE, Kamwendo D, Hoffman IF, Keating M, Brown LB, Ndalama B, Fiscus SA, Congdon S, Cohen MS, Miller WC. Detection of acute HIV infection: a field evaluation of the determine® HIV-1/2 Ag/Ab combo test. J Infect Dis. 2012 Feb 15;205(4):528–34.

24. Faraoni S, Rocchetti A, Gotta F, Ruggiero T, Orofino G, Bonora S, Ghisetti V. Evaluation of a rapid antigen and antibody combination test in acute HIV infection. J Clin Virol. 2013 May;57(1):84–7.

25. Brauer M, De Villiers JC, Mayaphi SH. Evaluation of the Determine™ fourth generation HIV rapid assay. J Virol Methods. 2013 Apr;189(1):180–3.

26. Kawahata T, Nagashima M, Sadamasu K, Kojima Y, Mori H. [Evaluation of an immunochromatographic fourth generation test for the rapid diagnosis of acute HIV infection]. Kansenshogaku Zasshi. 2013 Jul;87(4):431–4. Japanese

27. Pilcher CD, Louie B, Facente S, Keating S, Hackett J Jr, Vallari A, Hall C, Dowling T, Busch MP, Klausner JD, Hecht FM, Liska S, Pandori MW. Performance of rapid point-of-care and laboratory tests for acute and established HIV infection in San Francisco. PLoS One. 2013 Dec 12;8(12):e80629.

28. Conway DP, Holt M, McNulty A, Couldwell DL, Smith DE, Davies SC, Cunningham P, Keen P, Guy R; Sydney Rapid HIV Test Study. Multi-centre evaluation of the Determine HIV Combo assay when used for point of care testing in a high risk clinic-based population. PLoS One. 2014 Apr 8;9(4):e94062.

29. Duong YT, Mavengere Y, Patel H, Moore C, Manjengwa J, Sibandze D, Rasberry C, Mlambo C, Li Z, Emel L, Bock N, Moore J, Nkambule R, Justman J, Reed J, Bicego G, Ellenberger DL, Nkengasong JN, Parekh BS. Poor performance of the determine HIV-1/2 Ag/Ab combo fourth-generation rapid test for detection of acute infections in a National Household Survey in Swaziland. J Clin Microbiol. 2014 Oct;52(10):3743–8.

30. Ottiger C, Huber AR. Comparison of the new Alere HIV Combo with Alere Determine HIV-1/2 Ag/Ab Combo in actute and primo and established HIV infections. Annals of Clinical and Laboratory Research. 2015 Vol 3 No 3:22.

31. Stekler JD, Ure G, O’Neal JD, Lane A, Swanson F, Maenza J, Stevens C, Coombs RW, Dragavon J, Swenson PD, Golden MR. Performance of Determine Combo and other point-of-care HIV tests among Seattle MSM. J Clin Virol. 2016 Mar;76:8–13.

32. Smit E, Atherton C, Osman H. Determine HIV-1/2 Ab/Ag Combo lacks sensitivity in detecting early HIV infections. European Society for Clinical Virology winter meeting. London, 13–15 January 2011.

33. Delaugerre C, Antoni G, Mahjoub N, Pialoux G, Cua E, Pasquet A, Hall N, Tremblay C, Cotte L, Capitant C, Chaix ML, Meyer L, Molina JM; IPERGAY Study Group. Assessment of HIV Screening Tests for Use in Preexposure Prophylaxis Programs. J Infect Dis. 2017 Aug 1;216(3):382–386.

34. Fitzgerald N, Cross M, O’Shea S, Fox J. Diagnosing acute HIV infection at point of care: a retrospective analysis of the sensitivity and specificity of a fourth-generation point-of-care test for detection of HIV core protein p24. Sex Transm Infect. 2017 Mar;93(2):100–101.

35. Fransen K, de Baetselier I, Rammutla E, Ahmed K, Owino F, Agingu W, Venter G, Deese J, Van Damme L, Crucitti T; FEMPrEP study group. Performance of serological and molecular tests within acute HIV infection. J Clin Virol. 2017 Aug;93:81–84.

36. Livant E, Heaps A, Kelly C, Maharaj R, Samsunder N, Nhlangulela L, Karugaba P, Panchia R, Marrazzo J, Chirenje ZM, Parikh UM; VOICE Study Team. The fourth generation Alere^TM^ HIV Combo rapid test improves detection of acute infection in MTN-003 (VOICE) samples. J Clin Virol. 2017 Sep;94:15–21.

37. Masciotra S, Luo W, Westheimer E, Cohen SE, Gay CL, Hall L, Pan Y, Peters PJ, Owen SM. Performance evaluation of the FDA-approved Determine™ HIV-1/2 Ag/Ab Combo assay using plasma and whole blood specimens. J Clin Virol. 2017 Jun;91:95–100.

38. Stafylis C, Klausner JD. Evaluation of two 4th generation point-of-care assays for the detection of Human Immunodeficiency Virus infection. PLoS One. 2017 Aug 28;12(8):e0183944.

39. Parker MM, Bennett SB, Sullivan TJ, Fordan S, Wesolowski LG, Wroblewski K, Gaynor AM. Performance of the Alere Determine™ HIV-1/2 Ag/Ab Combo Rapid Test with algorithm-defined acute HIV-1 infection specimens. J Clin Virol. 2018 Jul;104:89–91.

40. van Tienen C, Rugebregt S, Scherbeijn S, Götz H, Geurts van Kessel C. The performance of the Alere HIV combo point-of-care test on stored serum samples; useful for detection of early HIV-1 infections? Sex Transm Infect. 2018 Aug;94(5):331–333.

41. Chavez PR, Bradley HM, Wesolowski LG, Violette LR, Katz DA, Niemann LA, McMahan VM, McDougal S, Cornelius-Hudson AM, Ethridge SF, Stekler JD, Delaney KP. Performance evaluation of four point-of-care HIV tests using unprocessed specimens. J Clin Virol. 2020 Mar;124:104282

42. Wratil PR, Rabenau HF, Eberle J, Stern M, Münchhoff M, Friedrichs I, Stürmer M, Berger A, Kuttner-May S, Münstermann D, Lucht A, Meixenberger K, Bannert N, Keppler OT. Comparative multi-assay evaluation of Determine™ HIV-1/2 Ag/Ab Combo rapid diagnostic tests in acute and chronic HIV infection. Med Microbiol Immunol. 2020 Apr;209(2):139–150.

43. Kerschberger B, Aung A, Mpala Q, Ntshalintshali N, Mamba C, Schomaker M, Tombo ML, Maphalala G, Sibandze D, Dube L, Kashangura R, Mthethwa-Hleza S, Telnov A, Tour R, Gonzalez A, Calmy A, Ciglenecki I. Predicting, Diagnosing, and Treating Acute and Early HIV Infection in a Public Sector Facility in Eswatini. J Acquir Immune Defic Syndr. 2021 Dec 15;88(5):506–517.

44. Sirivichayakul S, Pankam T, Pattanachaiwit S, Phancharoen K, Barisri N, Areeyolwattana S, Phanuphak N, Phanuphak P. Ability of Alere™ HIV Combo to diagnose acute HIV infection is based mainly on HIV-1 p24 antigen detection. Asian Pac J Allergy Immunol. 2021 Oct 31.

45. Guiraud V, Naizet A, Khan H, Benhafoun G, Hernandez P, Piccin L, Pichon A, Leng AL, Yousfi L, Gautheret-Dejean A. Fourth-Generation HIV Rapid Tests: Enhanced Sensitivity and Reduced Diagnostic Window for HIV-1 Primary Infection Screening. J Med Virol. 2024 Nov;96(11):e70044.

46. Manjate A, Nilsson C, Axelsson M, Lindroth S, Sirbu D, Sacarlal J, Andersson S, Unemo M. Laboratory-based evaluation of the 4th-generation AlereTM HIV Combo rapid point-of-care test. PLoS One. 2024 Feb 23;19(2):e0298912.

47. Ciglenecki I, Ntshalintshali N, Mukooza E, Lekelem S, Mavimbela M, Dlamini S, Dube L, Mabuza N, Haile M, Ellman T, Flores A, Keiser O, Matse S, de la Tour R, Calmy A, Kerschberger B. Low sensitivity of the fourth-generation antigen/antibody HIV rapid diagnostic test Determine™ HIV Early Detect for detection of acute HIV infection at the point of care in rural Eswatini: a diagnostic accuracy study. J Int AIDS Soc. 2025;28(7):e26517.

48. Taegtmeyer M, MacPherson P, Jones K, Hopkins M, Moorcroft J, Lalloo DG, Chawla A. Programmatic evaluation of a combined antigen and antibody test for rapid HIV diagnosis in a community and sexual health clinic screening programme. PLoS One. 2011;6(11):e28019.

49. O’Neal JD, Golden MR, Branson BM, Stekler JD. HIV nucleic acid amplification testing versus rapid testing: it is worth the wait. Testing preferences of men who have sex with men. J Acquir Immune Defic Syndr. 2012 Aug 1;60(4):e117–20.

50. Meulbroek M, Ditzel E, Saz J, Taboada H, Pérez F, Pérez A, Carrillo A, Font G, Marazzi G, Uya J, Cabrero J, Ingrami M, Marín R, Coll J, Pujol F. BCN Checkpoint, a community-based centre for men who have sex with men in Barcelona, Catalonia, Spain, shows high efficiency in HIV detection and linkage to care. HIV Med. 2013 Oct;14 Suppl 3:25–8.

51. Eu B, Roth N, Stoové M, O’Reilly M, Clarke E. Rapid HIV testing increases the rate of HIV detection in men who have sex with men: using rapid HIV testing in a primary care clinic. Sex Health. 2014 Mar;11(1):89–90.

52. Wouters K, Fransen K, Beelaert G, Kenyon C, Platteau T, Van Ghyseghem C, Collier I, Buyze J, Florence E. Use of rapid HIV testing in a low threshold centre in Antwerp, Belgium, 2007-2012. Int J STD AIDS. 2014 Nov;25(13):936–42.

53. Yang M, Prestage G, Maycock B, Brown G, de Wit J, McKechnie M, Guy R, Keen P, Fairley CK, Zablotska IB. The acceptability of different HIV testing approaches: cross-sectional study among GMSM in Australia. Sex Transm Infect. 2014 Dec;90(8):592–5.

54. Mayer K, Tomassilli J, Gelman M, Altfeld M, Allen T. HIV self-testing among men who have sex with men (MSM): acceptability of biweekly finger pricks to perform the alere determineTM HIV-1/2 Ag/Ab assay; AIDS 2014 -XX International AIDS Conference; July 2014; Melbourne, Australia.

55. Conway DP, Guy R, Davies SC, Couldwell DL, McNulty A, Smith DE, Keen P, Cunningham P, Holt M; Sydney Rapid HIV Test Study. Rapid HIV Testing Is Highly Acceptable and Preferred among High-Risk Gay And Bisexual Men after Implementation in Sydney Sexual Health Clinics. PLoS One. 2015 Apr 21;10(4):e0123814.

56. Conway DP, Guy R, McNulty A, Couldwell DL, Davies SC, Smith DE, Keen P, Cunningham P, Holt M; Sydney Rapid HIV Test Study. Effect of testing experience and profession on provider acceptability of rapid HIV testing after implementation in public sexual health clinics in Sydney. HIV Med. 2015 May;16(5):280–7.

57. Hoenigl M, Graff-Zivin J, Little SJ. Costs per Diagnosis of Acute HIV Infection in Community-based Screening Strategies: A Comparative Analysis of Four Screening Algorithms. Clin Infect Dis. 2016 Feb 15;62(4):501–511.

58. Hoenigl M, Chaillon A, Mehta SR, Smith DM, Graff-Zivin J, Little SJ. Screening for acute HIV infection in community-based settings: Cost-effectiveness and impact on transmissions. J Infect. 2016 Nov;73(5):476–484.

59. Alibayeva KO, Saparbekov MK, Baiserkin BS, Abishev AT, Tazhibaeva GH, Kasymbekova SZ. Study of the possibility of introduction of Kazakhstan NGO-based rapid HIV testing procedures. HIV AIDS (Auckl). 2019 Sep 11;11:219–227.

60. Green Kimberley. 2019. Detecting acute HIV infection to accelerate epidemic control. 2019. PATH, USAID, PEPFAR. Available at: https://www.whitehatcom.com/Alere/K_Green_HIV_4th_gen_V2_112019.pdf

61. Mullens AB, Duyker J, Brownlow C, Lemoire J, Daken K, Gow J. Point-of-care testing (POCT) for HIV/STI targeting MSM in regional Australia at community ‘beat’ locations. BMC Health Serv Res. 2019 Feb 2;19(1):93.

62. Frola CE, Zalazar V, Cardozo N, Vázquez ML, Arístegui I, Lucas M, Gun A, Cahn P, Sued O. Home-based HIV testing: Using different strategies among transgender women in Argentina. PLoS One. 2020 Mar 19;15(3):e0230429.

63. Martin EG, Cornett J, Mohammed DY, Salaru G. Rapid Testing Algorithm Performance in a Low-Prevalence Environment. Sex Transm Dis. 2020 May;47(5S Suppl 1):S35–S40.

64. Saz J, Dalmau-Bueno A, Meulbroek M, Pujol F, Coll J, Herraiz-Tomey Á, Pérez F, Marazzi G, Taboada H, Culqui DR, Caylà JA; BCN Checkpoint Working Group. Use of fourth-generation rapid combined antigen and antibody diagnostic tests for the detection of acute HIV infection in a community centre for men who have sex with men, between 2016 and 2019. PLoS One. 2021 Jul 27;16(7):e0255065. doi: 10.1371/journal.pone.0255065. Erratum in: PLoS One. 2021 Oct 11;16(10):e0258613.

65. Williams OD, Dean JA, Crothers A, Gilks CF, Gow J. Economic evaluation of alternative testing regimes and settings to detect undiagnosed HIV in Australia. BMC Health Serv Res. 2021 Jan 7;21(1):30. doi: 10.1186/s12913-020-06040-5. PMID: 33413386; PMCID: PMC7789789.

66. Ferrer P, Bastias C, Beltrán C, Afani A. Diagnosis of HIV infection using mass community rapid testing in Santiago, Chile. J Clin Virol Plus. 2022;2:100064.

67. Muñoz-Chimeno M, Valencia J, Rodriguez-Recio A, Cuevas G, Garcia-Lugo A, Manzano S, Rodriguez-Paredes V, Fernandez B, Morago L, Casado C, Avellón A, Ryan P. HCV, HIV AND HBV rapid test diagnosis in non-clinical outreach settings can be as accurate as conventional laboratory tests. Sci Rep. 2023 May 9;13(1):7554.

68. Ochwoto M, Matiang’i M, Machuki Onchieku N, Ndoria S, Matoke L, Otinga M, Zablon J, Mathebula E, Matoke-Muhia D. The feasibility and impact of deploying a four-tests panel at antenatal care in primary health care facilities of a developing country, Kenya. Front Public Health. 2024 Nov 25;12:1399612.

69. Rodriguez NM, Balian L, Kataki I, Tolliver C, Rivera-De Jesus J, Linnes JC. Stakeholder-engaged development of a rapid test for detection of acute HIV infection. Res Sq [Preprint]. 2024 Apr 19:rs.3.rs–4243639.

70. Kiptinness C, Kuo AP, Reedy AM, Johnson CC, Ngure K, Wagner AD, Ortblad KF. Examining the Use of HIV Self-Testing to Support PrEP Delivery: a Systematic Literature Review. Curr HIV/AIDS Rep. 2022 Oct;19(5):394–408.

71. Tieosapjaroen W, Williams E, Johnson CC, Baptista Da Silva C, Barr-DiChiara M, Ong JJ. Web Annex D. Should HIV testing services with rapid diagnostic tests and/or self-tests be used for initiation or continuation of injectable long-acting PrEP? In: Guidelines on lenacapavir for HIV prevention and testing strategies for long-acting injectable pre-exposure prophylaxis (PrEP). Geneva: World Health Organization; 2025. Available: 10.2471/B09479

72. van de Vijver D, van Kampen J, Wongsa A, Janamnuaysook R, Mesplede T, Matrajt L et al. Evaluating HIV testing strategies to reduce drug resistance during CAB-LA rollout in Thailand. A modelling study. 13th International AIDS Society Conference on HIV Science; 13–17 July; Kigali, Rwanda, 2025. Available: https://www.abstract-archive.org/Abstract/Share/91175

73. Cox SN, Wu L, Wittenauer R, Clark S, Roberts DA, Nwogu IB, Vitruk O, Kuo AP, Johnson C, Jamil MS, Sands A, Schaefer R, Kisia C, Baggaley R, Stekler JD, Akullian A, Sharma M. Impact of HIV self-testing for oral pre-exposure prophylaxis scale-up on drug resistance and HIV outcomes in western Kenya: a modelling study. Lancet HIV. 2024 Mar;11(3):e167–e175.

74. Oliveira Leite, L. Magno, F. Soares, D. Zeballos, L. Dezanet, O. Ferreira, M. Westin, D. Greco, A. Grangeiro, I. Dourado, The PrEP15-19 Choices study group. Equivalent performance of HIV Oral Fluid Self-Testing and Rapid Testing compared to Nucleic Acid Amplification Test in Screening Adolescents for Long-Acting Injectable Cabotegravir in Brazil. 13th International AIDS Society Conference on HIV Science; 13–17 July; Kigali, Rwanda, 2025. Available: https://www.abstract-archive.org/Abstract/Share/92394

75. Vermeulen M, van Schalkwyk C, Jacobs G, et al. The Impact of Early Antiretroviral Treatment (ART) for HIV on the Sensitivity of the Latest Generation of Blood Screening and Point of Care Assays. Viruses. 2022;14(7):1426.

76. Toolkit to optimize HIV testing algorithms. Geneva World Health Organization; 2021. Available: https://www.who.int/tools/optimizing-hiv-testing-algorithms-toolkit.

77. Smallwood M, Pant Pai N. Improving the Quality of Diagnostic Studies Evaluating Point of Care Tests for Acute HIV Infections: Problems and Recommendations. Diagnostics (Basel). 2017;7(1):13. Published 2017 Mar 4.

