## Supporting information for "Should antigen-antibody rapid diagnostic tests be used to detect acute HIV infection? A systematic review and meta-analysis of diagnostic performance"

### Appendix 1. PRISMA Checklist

#### Preferred Reported Items for Systematic Reviews and Meta-analyses (PRISMA)

| SECTION | # | CHECKLIST ITEM | PAGE # |
|---|---|---|---|
| **TITLE** | | | |
| Title | 1 | Identify the report as a systematic review, meta-analysis, or both. | 1 |
| **ABSTRACT** | | | |
| Structured abstract | 2 | Provide a structured summary including, as applicable: background; objectives; data sources; study eligibility criteria, participants, and interventions; study appraisal and synthesis methods; results; limitations; conclusions and implications of key findings; systematic review registration number. | 2 |
| **INTRODUCTION** | | | |
| Rationale | 3 | Describe the rationale for the review in the context of what is already known. | 3 |
| Objectives | 4 | Provide an explicit statement of questions being addressed with reference to participants, interventions, comparisons, outcomes, and study design (PICOS). | 3 |
| **METHODS** | | | |
| Protocol and registration | 5 | Indicate if a review protocol exists, if and where it can be accessed (e.g., Web address), and, if available, provide registration information including registration number. | 4 |
| Eligibility criteria | 6 | Specify study characteristics (e.g., PICOS, length of follow-up) and report characteristics (e.g., years considered, language, publication status) used as criteria for eligibility, giving rationale. | 4 |

| | | | |
|---|---|---|---|
| Information sources | 7 | Describe all information sources (e.g., databases with dates of coverage, contact with study authors to identify additional studies) in the search and date last searched. | 4 |
| Search | 8 | Present full electronic search strategy for at least one database, including any limits used, such that it could be repeated. | 4 |
| Study selection | 9 | State the process for selecting studies (i.e., screening, eligibility, included in systematic review, and, if applicable, included in the meta-analysis). | 4 |
| Data collection process | 10 | Describe method of data extraction from reports (e.g., piloted forms, independently, in duplicate) and any processes for obtaining and confirming data from investigators. | 5 |
| Data items | 11 | List and define all variables for which data were sought (e.g., PICOS, funding sources) and any assumptions and simplifications made. | 5 |
| Risk of bias in individual studies | 12 | Describe methods used for assessing risk of bias of individual studies (including specification of whether this was done at the study or outcome level), and how this information is to be used in any data synthesis. | 4 |
| Summary measures | 13 | State the principal summary measures (e.g., risk ratio, difference in means). | 5 |
| Synthesis of results | 14 | Describe the methods of handling data and combining results of studies, if done, including measures of consistency (e.g., $I^2$) for each meta-analysis. | 5 |
| Risk of bias across studies | 15 | Specify any assessment of risk of bias that may affect the cumulative evidence (e.g., publication bias, selective reporting within studies). | 4 |
| Additional analyses | 16 | Describe methods of additional analyses (e.g., sensitivity or subgroup analyses, meta-regression), if done, indicating which were pre-specified. | 5 |
| **RESULTS** | | | |
| Study selection | 17 | Give numbers of studies screened, assessed for eligibility, and included in the review, with reasons for exclusions at each stage, ideally with a flow diagram. | 5 |
| Study characteristics | 18 | For each study, present characteristics for which data were extracted (e.g., study size, PICOS, follow-up period) and provide the citations. | 6 |
| Risk of bias within | 19 | Present data on risk of bias of each study and, if available, | 7 |

| | | | |
|---|---|---|---|
| studies | | any outcome level assessment (see item 12). | |
| Results of individual studies | 20 | For all outcomes considered (benefits or harms), present, for each study: (a) simple summary data for each intervention group (b) effect estimates and confidence intervals, ideally with a forest plot. | 6 |
| Synthesis of results | 21 | Present results of each meta-analysis done, including confidence intervals and measures of consistency. | 8-9 |
| Risk of bias across studies | 22 | Present results of any assessment of risk of bias across studies (see Item 15). | 7 |
| Additional analysis | 23 | Give results of additional analyses, if done (e.g., sensitivity or subgroup analyses, meta-regression [see Item 16]). | 10-12 |
| **DISCUSSION** | | | |
| Summary of evidence | 24 | Summarize the main findings including the strength of evidence for each main outcome; consider their relevance to key groups (e.g., healthcare providers, users, and policy makers). | 13 |
| Limitations | 25 | Discuss limitations at study and outcome level (e.g., risk of bias), and at review-level (e.g., incomplete retrieval of identified research, reporting bias). | 14 |
| Conclusions | 26 | Provide a general interpretation of the results in the context of other evidence, and implications for future research. | 15 |
| **FUNDING** | | | |
| Funding | 27 | Describe sources of funding for the systematic review and other support (e.g., supply of data); role of funders for the systematic review. | 16 |

**Reference:** Page MJ, McKenzie JE, Bossuyt PM, et al. The PRISMA 2020 statement: an updated guideline for reporting systematic reviews. *BMJ*. 2021;372:n71. Published 2021 Mar 29. doi:10.1136/bmj.n71

### Appendix S2. Full search strategies

| | |
|---|---|
| **1. PubMed**<br><br>**Date of search:**<br><br>30 November 2024 | ("HIV"[MeSH Terms] AND ("diagnosis"[MeSH Subheading] OR "diagnosis"[All Fields] OR "screening"[All Fields] OR "mass screening"[MeSH Terms] OR ("mass"[All Fields] AND "screening"[All Fields]) OR "mass screening"[All Fields] OR "early detection of cancer"[MeSH Terms] OR ("early"[All Fields] AND "detection"[All Fields] AND "cancer"[All Fields]) OR "early detection of cancer"[All Fields] OR "screen"[All Fields] OR "screenings"[All Fields] OR "screened"[All Fields] OR "screens"[All Fields] OR "diagnosis"[MeSH Terms]) AND ("rapid diagnostic test"[Title/Abstract] OR "RDT"[Title/Abstract] OR "point-of-care"[Title/Abstract] OR "POC"[Title/Abstract] OR "test"[Title/Abstract] OR "algorithm"[Title/Abstract]) AND ("fourth-generation"[Title/Abstract] OR "4th generation"[Title/Abstract] OR "ag ab"[Title/Abstract] OR "antigen-antibody"[Title/Abstract] OR "Determine"[Title/Abstract] OR "Alere"[Title/Abstract]) AND ("acute hiv infection"[Title/Abstract] OR "AHI"[Title/Abstract] OR "early"[Title/Abstract] OR "recent"[Title/Abstract] OR "AEHI"[Title/Abstract])) AND (2010:2024[pdat]) |
| **2. EBSCO Medline** | MJ (HIV OR "acute HIV" OR "early HIV") AND (diagnosis OR screening) AND (sensitivity OR "diagnostic accuracy" OR "diagnostic performance" OR detection) AND ("fourth generation" OR "4th generation" OR Ag/Ab OR antigen-antibody) AND (test OR assay OR "rapid diagnostic test" OR RDT OR "point-of-care" OR POC) limited 01/01/2010 to 21/12/2024 |
| **3. ClinicalTrials-gov** | (HIV) AND (diagnosis OR screening OR early HIV detection) AND (rapid diagnostic test OR RDT OR point-of-care OR POC OR test OR algorithm) AND (fourth-generation OR 4th generation OR Ag/Ab OR antigen-antibody OR Determine OR Alere) AND |

| | |
|---|---|
| | (acute HIV infection OR AHI OR early OR recent OR AEHI) |
| **4. Cochrane Library** | HIV AND ("diagnostic accuracy" or sensitivity) AND (assay OR test OR "rapid diagnostic test" OR fourth-generation OR Ag7Ab OR antigen-antibody OR "Alere HIV Combo" OR "point of care" OR "Determine HIV-1/2 Ag/Ab Combo" OR "Determine Early Detect" |
| **5. Scopus** | (HIV OR "acute HIV" OR "early HIV") AND (diagnosis or screening) AND (sensitivity OR "diagnostic accuracy" OR "diagnostic performance" OR detection) AND (fourth-generation OR $4^{th}$ generation OR Ag/Ab OR antigen-antibody) AND (test OR assay OR "rapid diagnostic test" OR RDT OR "point of care") restricted 2010 to 2025 |
| **6. Web of Science** | ((((((AB=(Acute HIV)) OR AB=(HIV)) OR AB=(early HIV)) AND AB=(diagnosis OR screening)) AND AB=(sensitivity specificity OR diagnostic accuracy OR diagnostic performance OR detection)) AND AB=(fourth generation OR 4th generation OR Ag Ab OR antigen antibody)) AND AB=(test OR assay OR rapid diagnostic test OR RDT or point of care OR POC). Timespan: 2006-01-01 until 2025-01-01 |

### Table S1. Outcomes reported by each study

| Count | Study, Year | Country | WHO Region | Diagnostic Accuracy | | Patient-level impact outcomes | | | | | Broadeer implementation outcomes | | | | | |
|---|---|---|---|---|---|---|---|---|---|---|---|---|---|---|---|---|
| | | | | Diagnostic Accuracy | Nondiagnostic | Test Uptake | Positivity | Ag positivity | Link to confirmation | Link to ART | Feasibiliy | Acceptability | Values/Preferences | Social harms | Resource use | Barriers and facilitarors |
| 1 | Beelaert, 2010 | Belgium | EURO | X | | | | | | | | | | | | |
| 2 | Fox, 2011 | UK | EURO | X | | | | | | | | | | | | |
| 3 | Naylor, 2011 | UK | EURO | X | | | | | | | | | | | | |
| 4 | Taegmeyer, 2011 | UK | EURO | | X | | X | X | X | X | X | | | X | | X |
| 5 | Kilember, 2012 | Rwanda, Zambia | AFRO | X | | | | | | | | | | | | |
| 6 | Laperche, 2012 | France | EURO | X | | | | | | | | | | | | |
| 7 | Patel, 2012 | USA | AMRO | X | | | | | | | | | | | | |
| 8 | Rosenberg, 2012 | Malawi | AFRO | X | | | | | | | | | | | | |
| 9 | O'Neil, 2012 | USA | AMRO | | X | | | | | | | | X | | | X |
| 10 | Brauer, 2013 | South Africa | AFRO | X | | | | | | | | | | | | |
| 11 | Faraoni, 2013 | Italy | EURO | X | | | | | | | | | | | | |
| 12 | Kahawata, 2013 | Japan | WPRO | X | | | | | | | | | | | | |
| 13 | Pilcher, 2013 | USA | AMRO | X | | | | | | | | | | | | |
| 14 | Melbroek, 2013 | Spain | EURO | | X | | X | | | X | X | X | | | | X |
| 15 | Conway, 2014 | Australia | WPRO | X | | | | | | | | | | X | | |
| 16 | Duong, 2014 | Eswatini | AFRO | X | | | | | | | | | | | | |
| 17 | Eu, 2014 | Australia | WPRO | | X | X | X | X | X | | X | X | X | | | X |
| 18 | Wouters, 2014 | Belgium | EURO | | X | X | | | | X | X | X | | X | | X |
| 19 | Yang, 2014 | Australia | WPRO | | X | | | | | | | X | X | | | X |
| 20 | Mayer, 2014 | USA | AMRO | | X | | X | | X | X | X | X | X | | | X |
| 21 | Conway, 2015 | Australia | WPRO | | X | | | | | | | X | X | | | X |
| 22 | Conway, 2015 | Australia | WPRO | | X | | | | | | | X | X | | | X |
| 23 | Ottiger, 2015 | Switzerand | EURO | X | | | | | | | | | | | | |
| 24 | Hoengl, 2016 | USA | AMRO | | X | | | | | | | | | | X | |
| 25 | Hoengl, 2016 | USA | AMRO | | X | | | | | | | | | | X | |
| 26 | Smit, 2016 | UK | EURO | X | | | | | | | | | | | | |
| 27 | Stekler, 2016 | USA | AMRO | X | | | | | | | | | | | | |
| 28 | Delaugerre, 2017 | France | EURO | X | | | | | | | | | | | | |
| 29 | Fitzgerald, 2017 | UK | EURO | X | | | | | | | | | | | | |
| 30 | Fransen, 2017 | Belgium | EURO | X | | | | | | | | | | | | |
| 31 | Livant, 2017 | SA. Uganda, Zim | AFRO | X | | | | | | | | | | | | |
| 32 | Masciotra, 2017 | USA | AMRO | X | | | | | | | | | | | | |
| 33 | Stafylis, 2017 | USA | AMRO | X | | | | | | | | | | | | |
| 34 | Parker, 2018 | USA | AMRO | X | | | | | | | | | | | | |
| 35 | van Tienen, 2018 | The Netherlands | EURO | X | | | | | | | | | | | | |
| 36 | Alivayeva, 2019 | Kazakhstan | EURO | | X | | | | | | X | | | X | | X |
| 37 | Green, 2019 | Vietnam | WPRO | | X | | X | X | X | X | X | | | | | X |
| 38 | Mullens, 2019 | Australia | WPRO | | X | | | | | | | X | X | | | X |
| 39 | Chavez, 2020 | USA | AMRO | X | | | | | | | | | | | | |
| 40 | Frola, 2020 | Argentina | AMRO | | X | X | X | X | X | | X | | | | | X |
| 41 | Martin, 2020 | USA | AMRO | | X | | X | X | X | X | X | | | X | | X |
| 42 | Wratil, 2020 | Germany | EURO | X | | | | | | | | | | | | |
| 43 | Kerschberger, 2021 | Eswatini | AFRO | X | X | | X | X | X | X | X | | | | | X |
| 44 | Saz, 2021 | Spain | EURO | | X | | X | X | X | X | X | | | X | | |
| 45 | Sirivichayakul | Thailand | SEARO | X | | | | | | | | | | | | |
| 46 | Williams, 2021 | Australia | WPRO | | X | | | | | | | | | | X | |
| 47 | Ferrer, 2022 | Chile | AMRO | | X | | | | | | X | | | | | X |
| 48 | Munoz-Cimmero, 2023 | Spain | EURO | | X | X | X | X | X | X | X | X | | | | X |
| 49 | Guiraud, 2024 | France | EURO | X | | | | | | | | | | | | |
| 50 | Manjate, 2024 | Mozambique | AFRO | X | | | | | | | | | | | | X |
| 51 | Ochwoto, 2024 | Kenya | AFRO | | X | | X | X | | | X | X | X | | | X |
| 52 | Rodriguez, 2024 | USA | AMRO | X | X | | | | | | | | X | | | |
| 53 | Ciglenecki, 2025 | Eswatini | AFRO | X | | | | | | | | | | | | |
| | TOTAL COUNT | | | 31 | 23 | 4 | 11 | 9 | 8 | 9 | 14 | 10 | 9 | 6 | 3 | 19 |

### Table S2. RDT brands and testing operators evaluated in each study

| Study ID | Study setting | Test operator | Sample size | Specimen type | Test brand name | Manufacturer |
|---|---|---|---|---|---|---|
| Beelaert 2010 | Belgium, lab | Laboratory technician | 239 | Serum | Determine HIV-1/2 Ag/Ab Combo | Alere Medical, USA (now Abbott) |
| Fox 2011 | UK, lab | Laboratory technician | 26 | Serum | Determine HIV-1/2 Ag/Ab Combo | Alere Medical, USA (now Abbott) |
| Naylor 2011 | UK, hospital | Not reported | 5 | Fingerstick | Determine HIV-1/2 Ag/Ab Combo | Alere Medical, USA (now Abbott) |
| Kilembe 2012 | Rwanda, Zambia, lab | Laboratory Scientist | 59 | Plasma | Determine HIV-1/2 Ag/Ab Combo | Alere Medical, USA (now Abbott) |
| Laperche 2012 | France, lab | Laboratory staff | 20 | Plasma | Determine HIV-1/2 Ag/Ab Combo | Alere Medical, USA (now Abbott) |
| Patel 2012 | USA, clinics | Laboratorians | 33 | Plasma | Determine HIV-1/2 Ag/Ab Combo | Alere Medical, USA (now Abbott) |
| Rosenberg 2012 | Malawi, clinics | HIV counsellors | 846 | Whole blood | Determine HIV-1/2 Ag/Ab Combo | Alere Medical, USA (now Abbott) |
| Faraoni 2013 | Italy, hospital | Laboratory staff | 141 | Serum | Determine HIV-1/2 Ag/Ab Combo | Alere Medical, USA (now Abbott) |
| Brauer 2013 | SA, lab | Laboratory staff | 36 | Serum | Determine HIV-1/2 Ag/Ab Combo | Alere Medical, USA (now Abbott) |
| Kahawata 2013 | Japan, lab | Laboratory staff | 25 | Plasma | Espline HIV Ag/Ab test | Fujirebio, Japan |

| | | | | | | |
|---|---|---|---|---|---|---|
| Pilcher 2013 | USA, lab | Laboratory staff | 138 | Plasma | Determine HIV-1/2 Ag/Ab Combo | Alere Medical, USA (now Abbott) |
| Conway 2014 | Australia, clinics | Doctors and nurses | 3154 | Fingerstick | Determine HIV-1/2 Ag/Ab Combo | Alere Medical, USA (now Abbott) |
| Duong 2014 | Eswatini, at-home testing | Well-trained nurses | 12370 | Fingerstick | Determine HIV-1/2 Ag/Ab Combo | Alere Medical, USA (now Abbott) |
| Ottiger 2015 | Switzerland, lab | Laboratory staff | 21 | Serum | Determine HIV-1/2 Ag/Ab Combo and Alere HIV Combo | Alere Medical, USA (now Abbott), Abbott Alere Japan |
| Stekler 2016 | USA, clinic | Laboratory staff | 1494 | Whole blood | Determine HIV-1/2 Ag/Ab Combo | Alere Medical, USA (now Abbott) |
| Smith 2016 | UK, clinic | Laboratory staff | 21 | Serum | Determine HIV-1/2 Ag/Ab Combo | Alere Medical, USA (now Abbott) |
| Delaugerre 2017 | France, clinic | Laboratory staff | 13 | Plasma | Alere HIV Combo | Abbott Alere, Japan |
| Fitzgerald 2017 | UK, lab | Laboratory staff | 69 | Serum | Alere HIV Combo | Abbott Alere, Japan |
| Fransen et al 2017 | Belgium, lab | Laboratory staff | 64 | Plasma | Determine HIV-1/2 Ag/Ab Combo and Alere HIV Combo and SD Bioline HIV Ag/Ab Combo | Alere Medical, USA (now Abbott), Alere Abbott, Japan, Standard Diagnostics Korea (now Abbott) |
| Livant 2017 | SA, Uganda, Zim, clinics | Laboratory staff | 215 | Plasma | Determine HIV-1/2 Ag/Ab Combo and | Alere Medical, USA Abbott Alere, Japan (now Abbott) |

| | | | | | | |
|---|---|---|---|---|---|---|
| | | | | | Alere HIV Combo | |
| Masciotra 2017 | USA, lab | Laboratory staff | 112 | Plasma | Determine HIV-1/2 Ag/Ab Combo | Alere Medical, USA (now Abbott) |
| Stafylis 2017 | USA, lab | Laboratory staff | 49 | Serum | Determine HIV-1/2 Ag/Ab Combo and SD Bioline HIV Ag/Ab Combo | Alere Medical, USA,<br><br>Standard Diagnostics Korea (now Abbott) |
| Parker 2018 | USA, lab | Laboratory staff | 125 | Serum | Determine HIV-1/2 Ag/Ab Combo | Alere Medical, USA (now Abbott) |
| van Tienen 2018 | Netherlands, lab | Laboratory staff | 23 | Serum | Alere HIV Combo | Abbott Alere, Japan |
| Chavez 2020 | USA, clinic | Not reported | 878 | Venous blood | Determine HIV-1/2 Ag/Ab Combo | Alere Medical, USA (now Abbott) |
| Wratil 2020 | Germany, lab | Laboratory staff | 21 | Plasma | Determine HIV-1/2 Ag/Ab Combo and Alere HIV Combo | Alere Medical, USA (now Abbott) |
| Kerschberger 2021 | Eswatini, outpatient centre | Laboratory technicians | 731<br>745 | Plasma, whole blood | Alere HIV Combo | Abbott Alere, Japan |
| Sirivichayakul 2021 | Thailand, lab | Laboratory staff | 50 | Plasma | Alere HIV Combo | Abbott Alere, Japan |
| Guiraud 2024 | France, lab | Laboratory staff | 32 | Serum | Determine Early Detect | Abbott Alere, Japan |
| Manjate 2024 | Mozambique, outpatient centre | Laboratory staff | 762 | Serum | Alere HIV Combo | Abbott Alere, Japan |
| Ciglenecki 2025 | Eswatini, HIV testing sites | HTS counsellors | 1163 | Fingerstick | Determine Early Detect | Abbott Alere, Japan |

**Table S3. characteristics of the 23 included studies assessing patient-level impact and implementation outcomes**

| Study ID | Country | Setting | Population | Testing modality | Reported outcomes | Comparator & Study design |
|---|---|---|---|---|---|---|
| Taegmeyer 2011 | UK | Community outreach and hospital-based STI clinics | KP: PWID, MSM, SW, asylum seekers | Community-based testing | Test Positivity, Ag Positivity, Linkage to Confirmatory Testing and Treatment, Feasibility, social harms, and barriers | No comparator<br><br>Cross-sectional |
| O'Neil 2012 | USA | STI clinics | KP: MSM, TG | Facility-based testing | Values and Preferences and barriers | No comparator<br>Cross-sectional |
| Melbroek 2013 | Spain | Community-based centre | KP: MSM | Community-based testing | Test Positivity, Linkage to Treatment, Feasibility, Acceptability, and barriers | 6-year retrospective analysis of data<br>Retrospective cross-sectional |
| Eu 2014 | Australia | Primary healthcare clinic | KP: MSM | Facility-based testing | Test Uptake, Test Positivity, Ag Positivity, Linkage to Confirmatory Testing, Feasibility, Values, Preferences, and barriers | 32-month retrospective HIV positivity data<br><br>Cross-sectional |
| Wouters 2014 | Belgium | Outpatient screening centre | MSM, Southern African migrants and others | Facility-based testing | Test Uptake, Test Positivity, Linkage to Treatment, Feasibility, Acceptability, social harm, and barriers | 5-year retrospective analysis data<br>Retrospective cross-sectional |
| Yang 2014 | Australia | Online survey distributed through peer- | KP: MSM | Not applicable | Acceptability, Values, Preferences, and barriers | No comparator |

| | | | | | | |
|---|---|---|---|---|---|---|
| | | referral networks | | | | Cross-sectional |
| Mayer 2014 | USA | Home-based self-testing | KP: MSM | Home-based self-testing | Test Positivity, Linkage to Confirmatory Testing and Treatment, Feasibility, Acceptability Values & Preferences, and barriers | No comparator<br><br>Prospective Cohort |
| Conway 2015 | Australia | STI clinics | KP: MSM, TG | Facility-based testing | Acceptability, Values and Preferences, and barriers | No comparator<br><br>Longitudinal survey |
| Conway 2015 | Australia | STI clinics | Providers: doctors, nurses | Facility-based testing | Acceptability, Values and Preferences, and barriers | No comparator<br><br>Longitudinal survey |
| Hoenigl 2016 | USA | Community-based centre | KP: MSM | Community-based testing | Cost-effectiveness of 4 different testing strategies | Routine NAT Ag/Ab EIA Ag/Ab RDT Ab testing<br><br>Retrospective cohort |
| Hoenigl 2016 | USA | Community-based centre | KP: MSM | Community-based testing | Cost-effectiveness and impact on transmission | Routine NAT Ag/Ab EIA Ag/Ab RDT Ab testing<br><br>Retrospective cohort |
| Alivayeva 2019 | Kazakhstan | NGO clinic | PLHIV, HTC clients, government HIV providers, NGOs | Community-based testing | Diagnostic Accuracy, Feasibility, Social harm, and barriers | No comparator Mixed-methods Cross-sectional |
| Green 2019 | Vietnam | Community-based centres | KP: MSM, PLHIV and their partners | Community-based testing | Test Positivity, Ag Positivity, Linkage to Confirmatory Testing and Treatment, Feasibility, and barriers | Conventional PNS vs Enhanced PNS<br><br>Prospective Cohort |

| | | | | | | |
|---|---|---|---|---|---|---|
| Mullens 2019 | Australia | Mobile van clinic | KP: MSM | Community-based testing | Acceptability, Values and Preferences, and barriers | No comparator Mixed methods pilot |
| Frola 2020 | Argentina | Home-based testing | KP: TG | Home-based testing | Test Uptake, Test Positivity, Ag Positivity, Linkage to Confirmatory Testing, Feasibility, and barriers | No comparator<br><br>Cross-sectional |
| Martin 2020 | USA | CLIA-waived screening sites | General population | Facility-based testing | Test Positivity, Ag Positivity, Linkage to Confirmatory Testing and Treatment, Feasibility, social harm, and barriers | Rapid Testing Algorithm vs CDC Laboratory Algorithm<br><br>Retrospective cohort and DTA |
| Kerschberger 2021 | Eswatini | Outpatient ward of health centre | Individuals at high risk of acquiring HIV | Facility-based testing | Test Positivity, Ag Positivity, Linkage to Confirmatory Testing and Treatment, Feasibility, and barriers | Ag/Ab RDT vs VL Cross-sectional and prospective cohort |
| Saz 2021 | Spain | Community-based centre | KP: MSM | Community-based testing | Test Positivity, Ag Positivity, Linkage to Confirmatory Testing and Treatment, Feasibility and social harm | 3-year retrospective data analysis<br><br>Cross-sectional and DTA |
| Williams 2021 | Australia | Modelling across six testing settings | General population and MSM | Facility, Community, and home-based testing | Cost-effectiveness comparing 3 testing strategies | Conventional Parallel POC testing<br><br>Modelling study |
| Ferrer 2022 | Chile | University hospital | General population | Community-based testing | Diagnostic Accuracy, Feasibility, and barriers | No comparator Cross-sectional & DTA |

| | | | | | | |
|---|---|---|---|---|---|---|
| Muñoz-Chimeno 2023 | Spain | Mobile screening unit | Vulnerable population: PWUD, migrants, homeless people | Community-based testing with triplex RDT (HIV, HCV, HBV) | Test Uptake, Test Positivity, Ag Positivity, Linkage to Confirmatory Testing and Treatment, Feasibility, Acceptability, and barriers | No comparator Cross-sectional |
| Ochwoto 2024 | Kenya | Primary healthcare facilities | Pregnant women | Facility-based testing with 4-test panel (HIV, Syphilis, HBV, Malaria) | Test Positivity, Ag Positivity, Feasibility, Acceptability, Values & Preferences, and barriers | Retrospective analysis of participants' medical records<br><br>Cross-sectional |
| Rodriguez 2024 | USA | NGO | KP: PWUD, PLHIV | Community-based testing | Acceptability, Values and Preferences | No comparator Qualitative study |

### Table S4. Risk of bias assessment using the QUADAS-2 tool

#### Table 1. Study-level risk of bias and applicability across QUADAS-2 domains for diagnostic accuracy studies

| First author, year of publication and Ag/Ab RDT evaluated. | Patient Selection | Index test | Reference standard | Flow and timing |
|---|---|---|---|---|
| | Consecutive, random or retrospective selection? | Blinded to the results of reference test? | Blinded to the results of the index test? | Same sample? Same time? Same reference? Some patients excluded? |
| Beelaert 2010 FDA | High | Unclear | Low | Low |
| Fox 2011 FDA | High | Unclear | Low | Low |
| Naylor 2011 FDA | Low | Low | Low | Low |
| Kilembe 2012 FDA | High | Unclear | Low | Low |
| Laperche 2012 FDA | High | High | Low | Low |
| Patel 2012 FDA | High | Low | Low | Low |
| Rosenberg 2012 FDA | Low | Low | Low | Low |
| Faraoni 2013 FDA | High | Unclear | Low | Low |
| Brauer 2013 FDA | High | Unclear | Low | Low |
| Kahawata 2013 | High | Unclear | Low | Low |
| Pilcher 2013 FDA | High | High | Low | Low |
| Conway 2014 FDA | Low | Low | Low | Low |
| Duong 2014 FDA | Low | Low | Low | Low |
| Stekler 2016 FDA | Low | Low | Low | Low |
| Smit 2016 FDA | High | High | Low | Low |
| Masciotra 2017 FDA | Low | Low | Low | Low |
| Stafylis 2017 FDA | High | Unclear | Low | Low |
| Parker 2018 FDA | High | Unclear | Low | Low |
| Chavez 2020 FDA | Low | Low | Low | Low |
| Ottiger 2015 FDA & CE-marked | High | Unclear | Low | Low |
| Delaugerre 2017 CE-marked | High | Unclear | Low | Low |
| Fitzgerald 2017 CE-marked | High | Unclear | Low | Low |
| Fransen 2017 FDA & CE-marked | High | Unclear | Low | Low |
| Livant 2017 FDA & CE-marked | High | Unclear | Low | Low |
| van Tienen 2018 CE-marked | High | High | Low | Low |
| Wratil 2020 FDA & CE-marked | High | Unclear | Low | Low |

| | | | | |
|---|---|---|---|---|
| Kerschberger 2021 CE-marked | Low | Low | Low | Low |
| Sirivivhiyakul 2021 CE-marked | High | Unclear | Low | Low |
| Manjate 2024 CE-marked | Low | Low | Low | Low |
| Guiraud 2024 CE-marked | High | High | Low | Low |
| Ciglenecki 2025 CE-marked | Low | Low | Low | Low |

**Figure 1. Proportion of studies with low, high or unclear risk of bias**

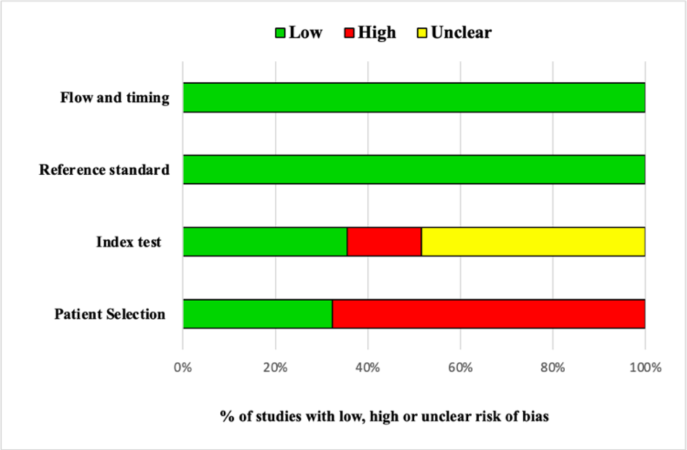


**Figure 2. Proportion of studies with low, high or unclear concerns regarding applicability**

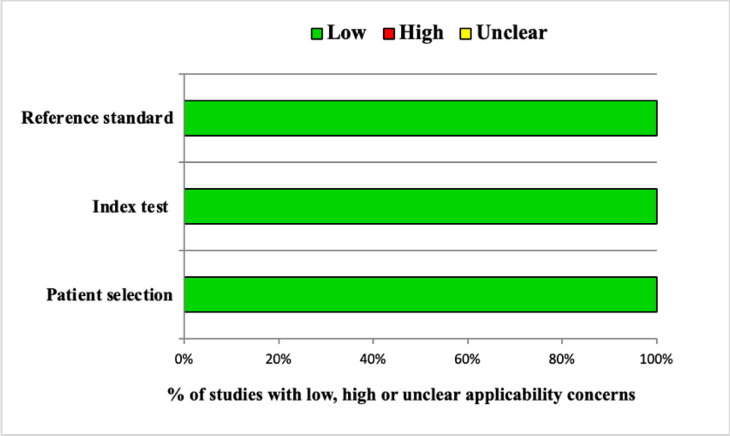

**Table S5. Risk of bias assessment using the JBI Critical Appraisal tool**

| Study Name | Taegmeyer 2011 | O'Neil 2012 | Melbroek 2013 | Eu 2014 | Wouters 2014 | Yang 2014 | Conway 2015 |
|---|---|---|---|---|---|---|---|
| 1. Were the criteria for inclusion in the sample clearly defined? | Yes | Yes | Yes | Yes | Yes | Yes | Yes |
| 2. Were the study subjects and the setting described in detail? | Yes | Yes | Yes | Yes | Yes | Yes | Yes |
| 3. Was the exposure measured in a valid and reliable way? | Yes | Yes | Yes | Yes | Yes | Yes | Yes |
| 4. Were objective, standard criteria used for measurement of the condition? | Yes | Yes | Yes | Yes | Yes | Yes | Yes |
| 5. Were confounding factors identified? | NA | NA | NA | NA | Yes | Yes | NA |
| 6. Were strategies to deal with confounding factors stated? | NA | NA | NA | NA | Yes | Yes | NA |
| 7. Were the outcomes measured in a valid and reliable way? | Yes | Yes | Yes | Yes | Yes | Yes | Yes |
| 8. Was appropriate statistical analysis used? | Yes | Yes | Yes | Yes | Yes | Yes | Yes |

| Study Name | Conway 2015 | Frola 2020 | Martin 2020 | Saz 2021 | Ferrer 2022 | Muñoz 2023 |
|---|---|---|---|---|---|---|
| 1. Were the criteria for inclusion in the sample clearly defined? | Yes | Yes | Yes | Yes | Yes | Yes |
| 2. Were the study subjects and the setting described in detail? | Yes | Yes | Yes | Yes | Yes | Yes |

| | | | | | | |
|---|---|---|---|---|---|---|
| 3. Was the exposure measured in a valid and reliable way? | Yes | Yes | Yes | Yes | Yes | Yes |
| 4. Were objective, standard criteria used for measurement of the condition? | Yes | Yes | Yes | Yes | Yes | Yes |
| 5. Were confounding factors identified? | Yes | Yes | Yes | NA | NA | NA |
| 6. Were strategies to deal with confounding factors stated? | Yes | Yes | Yes | NA | NA | NA |
| 7. Were the outcomes measured in a valid and reliable way? | Yes | Yes | Yes | Yes | Yes | Yes |
| 8. Was appropriate statistical analysis used? | Yes | Yes | Yes | Yes | Yes | Yes |

### Table S5. Risk of bias assessment using the ROBINS-I tool

| Study | Bias due to confounding | Bias in selection of participants into the study | Bias in classification of interventions | Bias due to deviations from intended inteventions | Bias due to missing data | Bias in measurement of outcomes | Bias in the selection of the reported result | Overall bias |
|---|---|---|---|---|---|---|---|---|
| Mayer 2014 | NI | Low risk | Low risk | Low risk | Low risk | NI | Low risk | Low risk |
| Green 2019 | NI | Low risk | Low risk | Low risk | Serious | Serious | Serious | High |
| Kerschberger 2021 | Low risk | Moderate | Low risk | Low risk | Low risk | Low risk | Low risk | Low risk |

**Appendix S3. Feasibility, acceptability, values & preferences, social harms, and resource use.**

### Feasibility

Of the fourteen included studies, ten (71%) addressed feasibility in well-resourced settings (UK [48], Australia [66], Belgium [52], Argentina [62], USA [54, 63], Spain [50, 64, 67], and Chile [66]), while four (29%) were conducted in resource-limited settings (Eswatini [43], Kazakhstan [59], Vietnam [60] and Kenya [68]). Most evaluations (62%) took place at the community level, including mobile units and home-based testing, with p24 Ag/Ab rapid tests performed by diverse providers (nurses, midwives, health assistants, counsellors, community providers, and laboratory technicians), primarily using whole blood specimens. The majority (77%) focused on key populations or individuals at high risk of HIV; the remaining studies involved the general population in the (USA, Chile) [63, 66] or pregnant women (Kenya) [68].

Three evaluations, two among the general population (USA screening sites [63], Chile mass community testing [66]) and one among key populations (UK, community testing [48]), concluded that p24 Ag/Ab RDT implementation was not feasible due to extremely low or absent acute HIV detection. However, these studies yielded important insights:

- The USA study established a fast-track pathway for p24-reactive results to accelerate confirmatory testing and linkage to care, while also noting inconsistent interpretation of results by operators during early field implementation, underscoring the need for robust training.
- Chile's programme validated 6 rapid tests prior to implementation of mass community testing, informing test selection and national policy.

Conversely, two programmes demonstrated high feasibility and effectiveness: a community-based MSM programme in Spain [64] and a health facility-based initiative in Eswatini (a high prevalence setting) [43]. Both achieved high testing uptake, linkage to confirmatory testing and rapid treatment initiation facilitated by near-point-of-care viral load systems. The Eswatini project also successfully implemented a risk-score algorithm to identify probable acute HIV infections. However, despite having on-site point-of-care viral load testing, confirmatory testing delays persisted due to limited testing capacity. Consequently, sameday treatment initiation was achieved in only 13% of patients, further constrained by low patient readiness to begin therapy.

Two evaluations, a mobile unit targeting key and vulnerable populations in Spain [67] and an antenatal clinic programme in Kenya [68], employed p24 Ag/Ab RDTs as part of integrated testing strategies. In Spain, testing included HIV, HBV, HCV by fingerstick blood, while the Kenyan programme used a 4-panel test for syphilis, malaria, HIV, HBV.

One study in Argentina [62] demonstrated the feasibility of a home-based HIV testing programme for transgender women, utilizing a multidisciplinary team and a screening algorithm (p24 Ag/Ab RDT followed by Ab-only RDT) with ELISA and pooled viral load testing for confirmation. The project achieved high testing uptake and linkage to care.

A study in Vietnam [60] evaluated a community-based index testing and enhanced partner notification services (PNS) programme for key populations and their partners. The intervention doubled HIV testing rates compared to routine PNS, though broader programmatic data are needed to assess the effectiveness of this model. Implementation challenges included delays in confirmatory testing due to weak linkages between the community testing sites and health facilities, which hindered rapid linkage to care.

### Acceptability

Ten observational studies evaluated client acceptability, with nine conducted in high-resource countries (Spain [50, 67], Australia [51, 53, 55, 61], Belgium [52], and USA [54, 69]) and one in a resource-limited setting (Kenya [68]). Most employed cross-sectional designs, with one qualitative study [69] and two mix-methods evaluations [61, 68]. Five studies additionally assessed provider acceptability (Belgium [52], Australia [55, 61], Kenya [68] and USA [69]).

Study populations included MSM [50, 51, 53–55], individuals at high risk of HIV [49, 69], key/vulnerable populations [67] and pregnant women [68]. Provider evaluations encompassed nurses [52, 68], doctors and nurses [56], peer test facilitators [61], and peer or community workers [54]. Implementation occurred across diverse settings: community centres [50], primary care clinics [51], screening sites [52], home-based testing [54], STI clinics [55], mobile units at the community [61, 67], antenatal clinics [68], testing and outreach units [69] and an online survey [53].

Quantified client acceptability rates were of 59% [55], 63% [53], 78% [61], 87% [52], 90% [54], and 98.6%, 99.7% and 100%, respectively [51, 67, 69]. Two studies from Spain and Kenya reported qualitatively high acceptability [50, 68]. Home-based self-testing with p24 Ag/Ab RDT was highly acceptable among MSM in the USA [54]. All five provider evaluations indicated high acceptability, though Australian clinicians noted greater physician acceptance due to the test's utility in medical decision-making [56].

### Values and Preferences

Nine observational studies assessed values and preferences regarding p24 Ag/Ab RDT use among key populations and pregnant women [49, 51, 53–56, 61, 68, 69], including one study evaluating HIV self-testing with p24 Ag/Ab RDT in home settings [54].

Benefits reported included ease-of-use and rapid results, convenience and reduced stress compared to laboratory-based testing, minimal discomfort with fingerstick blood sampling, and earlier detection of acute HIV infection versus Ab-only RDTs.

Two evaluations of integrated testing strategies in Spain and Kenya highlighted additional advantages including cost-saving through simultaneous multi-disease testing [67, 68]. High satisfaction among clients and providers, with most willing to recommend RDTs.

Client preferences consistently favoured nonclinical over clinical testing settings, fingerstick over venous blood sampling, increased testing frequency with rapid tests, and home-based self-testing (57% preferred for convenience and confidentiality [54]. For acute HIV infection, clients prioritized accuracy, preferring EIA/NAT over RDTs due to perceived reliability concerns [49].

Provided perspectives varied: Australian MSM indicated willingness to pay RDTs [70]. Kenyan nurses supported laboratory-based 4-panel test administration during the first ANC visits but cited concerns about costs and staffing constraints [68].

### Social harms and misdiagnosis

Social harm appeared rare, though data were limited. One feasibility study in Kazakhstan identified individual and social barriers to NGO-led rapid testing introduction, including fear of testing, self-stigmatization, social stigma and discrimination, as well as legal and financial constraints [59].

Five implementation studies documented false-positive results, which led to significant client anxiety during confirmation delays [43, 48, 52, 63, 64] and placed unnecessary burdens on laboratories [48, 63].

The primary causes of false positives included operator inexperience (particularly during initial rollout) [43, 63], misinterpretation of faint positive lines, and improper RDT storage conditions. Additionally, false positives were more likely in low-risk populations with very low prevalence [43].

### Resource use

Our review identified three costing analyses from high-resource settings (USA and Australia), with no cost evaluations available from resource-limited settings.

Two US studies in the USA examined cost-effectiveness of different HIV testing approaches for acute HIV detection at community level among MSM in San Diego, California. These compared: (1) antibody-only RDT with routine qualitative NAT (Early Detect), (2) p24 Ag/Ab EIA, (3) p24 Ag/Ab RDT, and (4) antibody-only testing.

The first study [57] analysed cost per test result and cost per AHI diagnosis, finding Early Detect cost-effective at HIV prevalence >0.4%, with p24 Ag/Ab EIA as the second most cost-effective option. While Early Detect showed optimal cost-effectiveness, the p24 Ag/Ab RDT algorithm had significantly lower overall costs. The study concluded that Ag/Ab RDTs could be cost-effective alternatives in settings currently using Ab-only RDTs.

The second study, a INSERT STUDY DESIGN compared X to Y [58] evaluated potential infections averted, demonstrated that while Early Detect was most cost-effective, it was also the most expensive option. Compared to antibody-only strategies, other p24 Ag/Ab algorithms showed favourable cost-effectiveness, averting 5.28, 4,22 and 2.78 more infections annually per strategy.

Lastly, an Australian DESIGN study [65] compared testing modalities (conventional EIA, parallel EIA/POC testing) and POC testing across general population and MSM. Results indicated Ag/Ab RDT as the most cost-effective option, even at 50% sensitivity in low prevalence settings.
